# A Randomized Safety and Feasibility Crossover Trial of two Mediterranean-Ketogenic Interventions in Individuals with Parkinson’s Disease

**DOI:** 10.1101/2025.04.01.25325057

**Authors:** Kira Tosefsky, Joyce S. T. Lam, Yolanda N. Wang, Shayan Keymanesh, Annie J. Kuan, Avril Metcalfe-Roach, Mihai S. Cirstea, Matthew A. Sacheli, B. Brett Finlay, Tamara R. Cohen, Silke Appel-Cresswell

## Abstract

**BACKGROUND:** Both Mediterranean and ketogenic diets have been proposed as nutritional interventions in Parkinson’s disease (PD). Combined approaches may offer maximal benefits.

**OBJECTIVE:** Assess the feasibility, safety and exploratory efficacy of two Mediterranean-ketogenic dietary interventions in individuals with PD (PwP).

**METHODS:** In this Phase II, random-order crossover study, PwP followed two 8-week dietary interventions, separated by an 8-week washout: 1) a high-fat, low-carbohydrate Mediterranean diet (MeDi-KD) and 2) a standard Mediterranean diet supplemented with medium chain triglycerides (MeDi-MCT).

**RESULTS:** Of 52 participants randomized, 48 started the trial. Forty-one (79%) participants completed at least one and 33 (63%) completed both intervention phases. There were no intervention-related serious adverse events, nor any significant changes in plasma lipid profiles. Seventy-three percent and 92% of participants reported deviating from the MeDi-KD and MeDi-MCT no more than a few times per month, respectively. Moderate Mediterranean Diet Adherence Scores of 6.7 (SD: 1.6) and 7.2 (SD: 2.3) were achieved during the MeDi-KD and MeDi-MCT, respectively, out of a maximum of 14. Fifty percent of participants were in nutritional ketosis (BHB >0.5 mM) at follow-up for the MeDi-KD, as compared with only 1 (3%) participant following the MeDi-MCT. MDS-UPDRS Part II and IV scores decreased by a mean of -1.4 (SD: 4.2; p=0.039) and -1.0 (SD: 3.0; p=0.044) points, respectively, following the MeDi-MCT.

**CONCLUSIONS:** Mediterranean-ketogenic interventions appear safe and potentially beneficial in PwP. Behavioral strategies to optimize adherence should be employed in future phase III trials.

**TRIAL REGISTRATION:** The trial was registered on ClinicalTrials.gov: NCT05469997.

**PLAIN LANGUAGE SUMMARY:** Many patients want to know what the best diet is to follow for their Parkinson’s disease. Previous studies have suggested benefits of both ketogenic diets, which are high in fats and low in carbohydrates, and Mediterranean-style diets. Combining these two approaches could offer maximal advantages, while reducing the challenges of following a strict ketogenic diet.

The aim of this study was to test whether so-called “Mediterranean-ketogenic” diets would be safe and practical for patients with Parkinson’s disease to follow. Early indicators of potential clinical benefit were also assessed. Fifty-two participants were randomly assigned to follow two types of Mediterranean-ketogenic diets for eight weeks each, with an eight-week break in between. The two interventions were: 1) A high-fat, low carbohydrate version of a Mediterranean diet (MeDi-KD) and 2) A Mediterranean diet administered with a ketogenic supplement, medium-chain triglyceride oil (MeDi-MCT).

Forty-eight people started the study, 41 completed at least one diet intervention phase and 33 completed both intervention phases. No serious side effects were reported. Although most participants felt they followed the diets closely, objective measures of adherence suggested room for improvement.

The MeDi-MCT diet led to small but significant improvements in patient-reported motor activities of daily living and motor complications. Longer-term studies are needed to validate these findings. Future trials should incorporate behavioral coaching techniques to better help participants follow the diets.

## INTRODUCTION

Parkinson’s disease (PD) is the second most common neurodegenerative disease,^1,2^ and fastest growing neurological disorder worldwide.^3,4^. Current PD treatments primarily target cardinal motor symptoms, with no therapies yet found to slow or halt disease progression, other than potentially exercise.^5,6^ In this context, nutritional interventions such as the ketogenic diet (KD) have gained increased attention as a potential complementary approach to target multiple disease pathways in tandem, with putative disease-modifying and symptomatic benefits.^7,8^

Interest in ketogenic interventions for PD originated from preclinical models, which demonstrated that ketone bodies can attenuate neurodegeneration by supplying an alternative fuel source to glucose for bioenergetically compromised nigral neurons.^9^ Simultaneously, ketone bodies have been shown to reduce disease-related mitochondrial dysfunction, oxidative stress and neuroinflammation.^9^ Supply of ketone bodies to the brain may be increased through adherence to a high-fat, low-carbohydrate KD (70-90% calories from fat, 2-10% from carbohydrates, and 8-20% from protein),^10^ or through consumption of ketogenic supplements such as medium-chain triglycerides (MCTs).^11,12^ Previous short-term pilot studies of ketogenic interventions in PD have reported variable levels of improvement across multiple symptom domains, including voice quality,^13^ exercise performance,^14^ anxiety,^15^ and self-reported non-motor,^15–18^ motor,^17,18^ and cognitive ^19^ symptoms.

While this preliminary evidence is encouraging, several important safety concerns have been raised regarding the long-term implementation of standard ketogenic regimens in persons with PD (PwP). Standard KDs may be associated with poor diet quality and micronutrient deficiencies,^20–22^ for which PwP are at elevated baseline risk.^23^ Standard KDs are often high in saturated fats, which may increase levels of cardiovascular risk biomarkers,^24–27^ and have been shown to increase the risk of major adverse cardiovascular events over long-term follow-up.^28^ KDs may induce fluid loss through increased urination, leading to hypotension and increased risk of falls, which is of particular concern in the elderly population to which many PD patients belong and postural hypotension due to PD.^29^ Weight loss, which has been associated with reduced health-related quality of life,^30^ and increased mortality in PD, is a common effect of KDs,^24,25^ and has been consistently observed in five of the six PD ketogenic trials wherein this was reported.^15–19^ Additionally, KDs have been associated with shifts in gut microbiota composition which may be specifically or especially unfavourable in the setting of PD,^31^ wherein gut dysbiosis may play a causative role in disease through interactions with the immune and enteric nervous systems.^32–35^

Combining principles from a KD with those of the Mediterranean diet represents one strategy to improve the safety profile of ketogenic regimens,^36,37^ particularly in the setting of PD wherein Mediterranean diets have been repeatedly associated with favourable outcomes.^38–46^ Relative to standard ketogenic meal plans, Mediterranean-ketogenic diets permit some increased carbohydrate consumption primarily in the form of non-starchy vegetables, thereby protecting against micronutrient insufficiency, providing nutritional fiber and promoting the growth of healthy gut flora.^47^ At the same time, these diets emphasize unsaturated fat sources, such as olive oil and fish, which may minimize cardiovascular risks associated with diets high in saturated fats.^37^

Various neurologic, metabolic and gastrointestinal health benefits of Mediterranean-ketogenic diets have been reported in mild cognitive impairment,^37,47–50^ and Alzheimer’s disease,^51^. However, only one study to date has focused on adherence to Mediterranean-ketogenic diets in community-dwelling older adults,^37^ which is of particular relevance given the substantial adherence challenges associated with intense carbohydrate restriction in trials of standard KDs.^21,52–54^ As an alternative Mediterranean-ketogenic approach,^55^ the comparative safety and acceptability of an MCT-supplemented Mediterranean diet without carbohydrate restriction remains to be evaluated.

In this “Mediterranean-Ketogenic Interventions in Parkinson’s Disease” (KIM) Trial, we describe the safety and feasibility outcomes from a phase II, random-order crossover trial of an 8-week Mediterranean-ketogenic (MeDi-KD) and 8-week MCT-supplemented Mediterranean (MeDi-MCT) diet in PwP, separated by an 8-week washout. We assess feasibility in terms of recruitment capability, retention and dietary adherence, and report on safety indicators including adverse events, changes in fasting lipid profiles, and weight loss. Changes in scores on the Movement Disorder Society-Unified Parkinson’s Disease Rating Scale (MDS-UPDRS) Parts I-IV,^56^ Montreal Cognitive Assessment (MoCA),^57^ and Parkinson’s Disease Cognitive Rating Scale (PD-CRS),^58^ were examined on an exploratory basis.

## METHODS

### 1. Participants and Recruitment

The study protocol was approved by the University of British Columbia Clinical Research Ethics Board (Protocol #: H21-03747) and was registered on clinicaltrials.gov (<u>NCT05469997</u>). Participants were recruited on a rolling basis from the patient database of a tertiary Movement Disorders Clinic in Vancouver, Canada or through self-referral. Potential participants were pre-screened for eligibility based on review of clinician notes in the electronic medical record, and/or on the basis of physician comments during weekly research meetings. Written consent was obtained from all participants prior to enrolment.

Functionally independent individuals aged 40-85 years, with mild-to-moderate PD without dementia (MoCA ≥21), who were on stable or no dopaminergic medication for ≥1 month, met the study’s principal inclusion criteria (**Supplemental File 1; Supplemental Table 1**).

Participants on warfarin were excluded due to potential adverse effects of diet-induced changes in vitamin K on warfarin efficacy,^59^ and those on insulin were excluded due to the risk of hypoglycemia.^60^ Individuals with inflammatory bowel disease and/or a recent history of probiotic, antibiotic or immunomodulatory medication usage were excluded due to potential interference with gut microbiome-related readouts,^61^ which will be reported in a separate publication.

Prospective participants were contacted via phone. Inclusion and exclusion criteria were reviewed during the initial phone call, after which interested participants were scheduled for consenting and first study visits. Final eligibility checks were conducted in person at the first study visit.

### 2. Randomization

Eligible participants were randomized to start on either the MeDi-KD or MeDi-MCT intervention using variable block sizes with a 1:1 ratio.

### 3. Dietary Intervention

At the first study visit, personalized nutritional guidance was provided to participants) during a 1-hour consultation with the study’s registered dietitian (YNW). Baseline dietary intakes were assessed using the Canadian Diet History Questionnaire II.^62^ All participants were provided with online access to recipes via the KetoDietCalculator website (https://www.ketodietcalculator.org/about/). Participants were assigned to one of four meal plans based on anthropometry-based estimations of caloric requirements using the Harris-Benedict equation: 1500 kcal/day, 1750 kcal/day, 2000 kcal/day, or 2250 kcal/day.^63^ If participants struggled to maintain their desired body weight for successive weeks, they were instructed by the dietitian to add one or more 200 kcal snacks to the baseline meal plan. Participants were provided gram scales to measure out ingredients. Coconut-derived MCT oil (Nutiva MCT oil (Nutiva Inc.); NPN: 80086912) was supplied to participants starting the MeDi-MCT intervention. All participants were shown how to use the Abbott Freestyle Precision Neo device for at-home weekly blood ketone self-monitoring. Participants were instructed to test their blood ketone levels 2 hours after their evening MCT dose (MeDi-MCT) or evening meal (MeDi-KD). Participants were supplied with study journals in which they recorded bowel movements, food intake on pseudo-random days once weekly, daily MCT oil intake (MeDi-MCT arm only), and any adverse side effects. Participants completed weekly 30-minute check-in phone calls with the dietitian throughout each intervention phase, wherein study journal entries were reviewed and individualized modifications made as needed. Care partners were invited to join all dietitian intake consultations and subsequent follow-ups.

In both intervention phases, the MeDi component emphasized minimally processed plant-based foods, nuts, seeds, fish, poultry, non-starchy vegetables, and olive oil, while limiting red meat, animal-based culinary fats, sweets, and carbonated beverages. In the MeDi-KD, carbohydrates were restricted to 4-10% of daily calories, with the remaining calories derived from lean proteins (10-20%) and (primarily unsaturated) fats (69-82%). The target ketogenic ratio (the ratio of fats to carbohydrates and protein by weight) was increased from 1:1 to 1.5:1 or 2:1 over the first two intervention weeks, depending on participants’ tolerance to the diet and ketone reading. As the Mediterranean diet does not explicitly provide specific guidance on macronutrient intake, participants in MeDi-MCT phase were asked to have food groups in proportions based on Canada’s Food Guide (1/2 of their plate fruit and vegetables, 1/4 protein, and 1/4 whole grain), while within each food group, the food selection is guided by the Mediterranean diet. During the MeDi-MCT phase, MCT doses were increased over the course of the first two weeks until one of the following thresholds were reached: 1) The dose was not tolerable, 2) the limit of the recommended dose on the supplements’ label was reached (i.e., ≤40 mL of MCT oil supplement/day), or 3) previously set target dose were reached (30ml/day) and ketosis (blood BHB levels >0.5 mM) was achieved. Participant handouts containing education and instructions for each diet are provided in **Supplemental File 2.**

### 4. Feasibility Assessment

We focused on three key domains of feasibility: recruitment capability, retention and dietary adherence.

1. Recruitment capability was assessed based on whether or not our target enrollment of 50 participants was met within 24 months of the study start date.
2. Retention rates were tracked through each study stage, with a prespecified 75% target completion rate. Reasons for withdrawal were documented in all cases where participants remained contactable.
3. Dietary adherence was tracked using a combination of once-weekly at-home measurements of blood beta-hydroxybutyrate concentration ([BHB]), pre-and post-intervention fasting plasma [BHB], pre- and post-intervention scores on the 14-item Mediterranean Diet Adherence Screener (MEDAS),^64^ and self-assessed adherence according to a 4-point scale administered at post-intervention visits. The MEDAS score awards points for olive oil, fruit, vegetable, legume, fish, nut, and red wine consumption as well as preference for white over red meat, while deducting points for consumption of animal fast, pastries and sugar-sweetened beverages.^64^ The 4-point scale asked participants how often they followed the diet, with the following answer options: 1) Followed the diet every single day 2) Deviated from the diet several times a month 3) Deviated from the diet more than one time a week 4) Could barely execute the diet. Elevated blood [BHB] was defined as 0.30 mM in the non-fasting, and 0.50 mM in the fasting state.^36,54,65,66^

Intervention acceptability was also assessed through semi-structured interviews, which will be analyzed qualitatively and reported in a separate publication.

### 5. Safety assessment

Participants were instructed to report adverse events to study team members at the time of occurrence and were asked about adverse effects at each check-in and study visit. All adverse events were reported to the Principal Investigator (SAC) and graded according to severity, persistence and probability of relatedness to diet. Fasting plasma lipid panels (total cholesterol, triglycerides, high-density lipoprotein cholesterol (HDL-C), and low-density lipoprotein cholesterol (LDL-C)) were obtained at pre- and post-intervention visits to monitor cardiovascular risk. Plasma lipid analysis was performed by the UBC Hospital Outpatient Laboratory.

### 6. Anthropometric Measurements

Weight and height were measured at the first study visit to calculate BMI, and weight was measured at each follow-up visit. Participants were discouraged from changing anti-Parkinsonian medications during the trial unless medically necessary and were instructed to inform the study team of any such changes during weekly check-in phone calls and follow-up study visits.

### 7. Clinical Assessment

The MDS-UPDRS, MoCA and Parkinson’s Disease Cognitive Rating Scale (PD-CRS) were assessed in the “on” state at each study visit by certified research personnel.^56–58^

### 8. Statistical analysis

All analyses were conducted in RStudio (Version 2024.12.1+563). Statistical significance was set a priori at p<0.05. Differences in baseline characteristics between randomization groups were compared using Wilcoxon rank sum tests, Pearson’s Chi-squared tests, Fisher’s exact tests and Wilcoxon rank sum exact tests dependent on variable type. Between- and within-intervention comparisons were performed using linear mixed effects models and Wilcoxon signed rank tests, respectively. Mixed effects models were adjusted for period-specific baselines and intervention sequence, with participant IDs included as a random effect. Reported results derive from available-case analyses. Linear associations between continuous variables were calculated using Pearson correlation. Relationships between ordinal variables were described using Spearman rank correlation. Statistical approaches were reviewed by a staff statistician.

## RESULTS

### 1. Recruitment

Sixty-eight participants were recruited to the study between April 2023 and April 2024, with the final participant completing the study in October 2024. **Figure 1** presents the flow of study participants according to the CONSORT guidelines. Fifty-two participants were randomized to start either the MeDi-KD or the MeDi-MCT intervention. Four of these participants withdrew before starting the first dietary intervention to which they were randomized, leaving a total of 48 participants having started one of the two diets.

**Figure 1.**
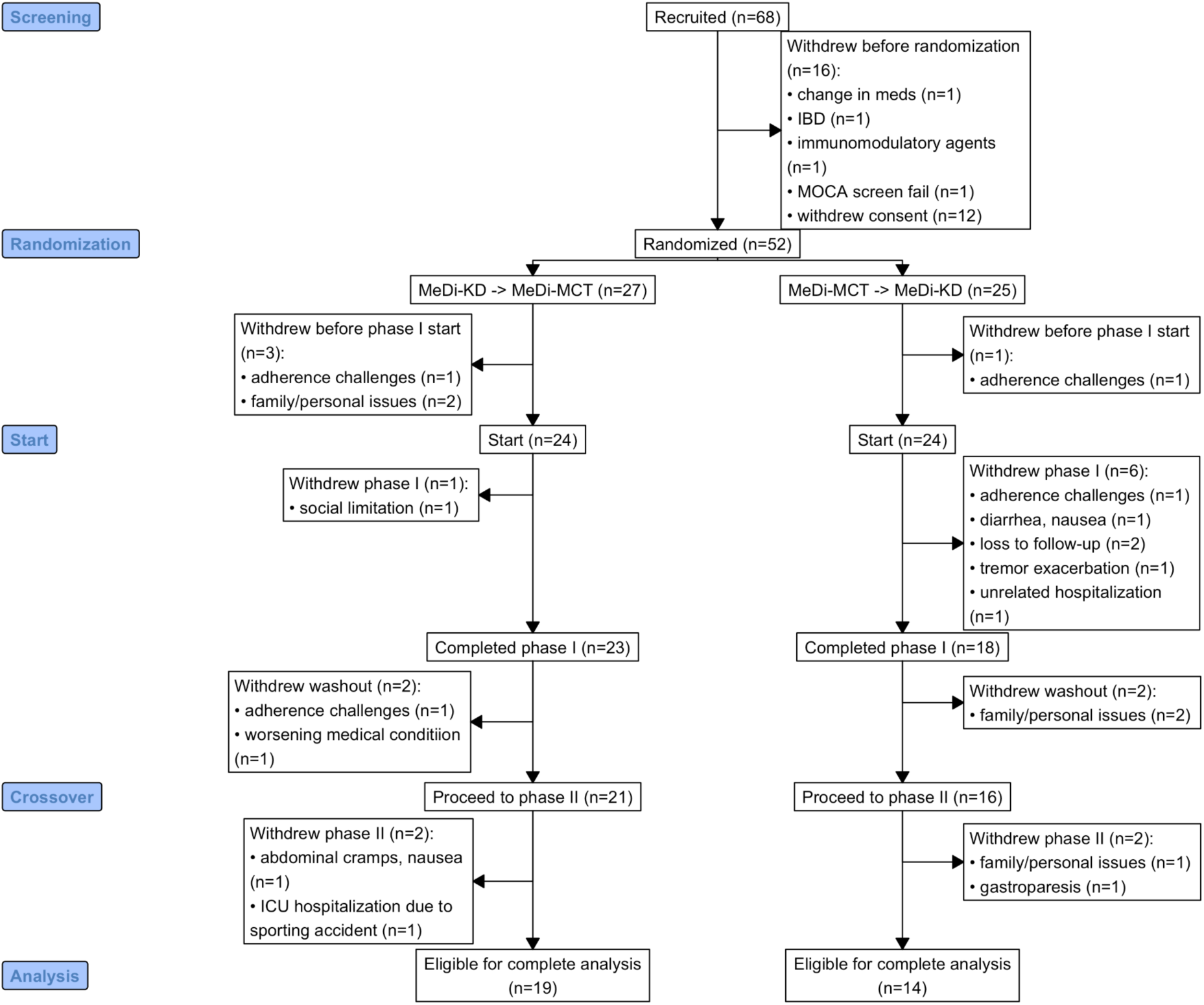
Consort diagram.

### 2. Baseline Characteristics

The median age of randomized participants was 68 (interquartile range [IQR]: 63-74), with a median disease duration of 7 (IQR: 4-10) years. Sixteen (31%) participants were female and 84% were Caucasian **(Table 1)**. Most participants had completed post-secondary education, with a median total of 18 (IQR: 16-20) years of education across the cohort. Eighty percent of participants were retired or not working. Most (76%) participants were married or in a long-term relationship, of whom 67% were living with a partner and 18% were living with children.

**Table 1.** Baseline Characteristics.

| Characteristic | Overall<br>N = 52<sup>1</sup> | MeDi-KD<br>N = 27<sup>1</sup> | MeDi-MCT<br>N = 25<sup>1</sup> | p-value<sup>2</sup> |
| --- | --- | --- | --- | --- |
| Disease Duration | 7 (4, 10) | 8 (4, 11) | 7 (4, 10) | 0.5 |
| Age | 68 (63, 74) | 71 (63, 75) | 67 (62, 69) | 0.10 |
| Sex |  |  |  | 0.4 |
| Female | 16 (31%) | 7 (26%) | 9 (36%) |  |
| Male | 36 (69%) | 20 (74%) | 16 (64%) |  |
| Ethnicity |  |  |  | 0.2 |
| Black/African | 1 (2.0%) | 0 (0%) | 1 (4.2%) |  |
| Caucasian | 41 (84%) | 22 (88%) | 19 (79%) |  |
| East Asian | 4 (8.2%) | 3 (12%) | 1 (4.2%) |  |
| South Asian | 3 (6.1%) | 0 (0%) | 3 (13%) |  |
| Years Education | 18 (16, 20) | 17 (16, 20) | 18 (17, 20) | 0.6 |
| BMI | 25.8 (22.3, 28.7) | 26.7 (22.6, 28.5) | 24.9 (22.3, 28.7) | 0.6 |
| Levodopa Equivalent Dose (mg/day) | 625 (375, 950) | 663 (550, 910) | 500 (10, 1,130) | 0.3 |
| Hoehn and Yahr Stage |  |  |  | 0.5 |
| 0 | 0 (0%) | 0 (0%) | 0 (0%) |  |
| I | 7 (13%) | 5 (19%) | 2 (8.0%) |  |
| II | 41 (79%) | 20 (74%) | 21 (84%) |  |
| III | 4 (7.7%) | 2 (7.4%) | 2 (8.0%) |  |
| Employed | 10 (20%) | 5 (20%) | 5 (21%) | >0.9 |
| Partnered | 37 (76%) | 18 (72%) | 19 (79%) | 0.6 |
| Married | 34 (67%) | 17 (65%) | 17 (68%) | 0.8 |
| MEDAS Score | 5.00 (4.00, 6.00) | 5.00 (4.00, 6.00) | 6.00 (4.00, 7.00) | 0.6 |
| MDS-UPDRS Part I | 10.5 (6.5, 14.0) | 11.0 (8.0, 14.0) | 10.0 (6.0, 13.0) | 0.5 |
| MDS-UPDRS Part II | 8.0 (5.5, 12.0) | 8.0 (6.0, 11.0) | 9.0 (4.0, 13.0) | 0.7 |
| MDS-UPDRS Part III | 26 (18, 36) | 25 (16, 34) | 26 (19, 37) | 0.5 |
| MDS-UPDRS Part IV | 4.0 (0.0, 7.0) | 4.0 (0.0, 7.0) | 4.0 (0.0, 7.0) | 0.9 |
| MoCA Total Score | 27.00 (25.00, 29.00) | 27.00 (26.00, 29.00) | 26.50 (25.00, 29.00) | 0.5 |
| PD-CRS Total Score | 91 (71, 106) | 92 (72, 106) | 87 (70, 105) | 0.8 |
| Constipation | 12 (23%) | 5 (19%) | 7 (28%) | 0.4 |
| Type II Diabetes | 2 (7.7%) | 0 (0%) | 2 (17%) | 0.2 |
| Dyslipidemia | 10 (29%) | 5 (28%) | 5 (31%) | >0.9 |
| Hypertension | 10 (31%) | 4 (25%) | 6 (38%) | 0.4 |
| Total energy intake (kcal/day) | 1,869 (1,336, 2,586) | 1,901 (1,330, 2,767) | 1,605 (1,341, 2,178) | 0.4 |
| % Calories from fat | 38 (35, 43) | 37 (35, 47) | 39 (32, 42) | 0.5 |
| % Calories from carbohydrates | 42.3 (39.4, 50.0) | 41.7 (37.5, 50.1) | 42.4 (41.3, 49.4) | 0.4 |

<sup>1</sup>Median (Q1, Q3); n (%)
<sup>2</sup>Wilcoxon rank sum test; Pearson's Chi-squared test; Fisher's exact test; Wilcoxon rank sum exact test

Forty-one (79%) participants had Hoehn and Yahr Stage 2 disease with a median MDS-UPDRS total score of 46 (IQR: 41-63) at the first study visit. The median levodopa equivalent dose was 625 (IQR: 375-950) mg/day. The median BMI was 25.8 (IQR: 22.3-28.7). Twelve (23%) participants experienced clinically significant constipation prior to starting the trial according to the ROME III module criteria.^67,68^

No participants were following a ketogenic diet or taking an MCT supplement at enrolment, as per the study’s exclusion criteria. At baseline, participant’s diets were generally not aligned with a Mediterranean pattern, with a median MEDAS score of 5 (IQR: 4-6) out of a maximum of 14.^69^

The two groups (those randomized to start on the MeDi-KD vs. MeDi-MCT intervention in phase I) were well balanced at baseline with respect to pertinent clinical and demographic variables, indicating that randomization was successfully achieved.

### 3. Retention

Forty-one (79%) participants completed at least one intervention, and 33 (63%) completed both intervention phases, falling short of our initial retention target of 75%. Seven of 16 (44%) female and 12 of 36 (33%) male participants withdrew (p=0.5). Study completion status did not differ based on randomization group (p=0.6). Study completion rate was higher (76%) among those with long-term partners or spouses as compared to participants who were single (33%) (p=0.027). Reasons for study withdrawal at each study stage are detailed in **Figure 1**.

### 4. Adherence

#### MEDAS Scores

Modest increases in MEDAS scores were seen following both the MeDi-KD (p=0.005) and MeDi-MCT (p=0.001) interventions **(Figure 2)** to a mean post-intervention score of 6.7 (standard deviation [SD]: 1.6) and 7.2 (SD: 2.3), respectively. No significant carryover effects in MEDAS scores were observed, as might have been expected if participants maintained a Mediterranean-style diet during the washout period. Examining MEDAS scores at each study visit according to previously established categories for weak, moderate and good adherence,^69^ we see that the majority of participants (76% on MeDi-KD; 54% on MeDi-MCT) were only able to achieve moderate adherence during either intervention phase, although 8 (23%) achieved good adherence during the MeDi-MCT intervention **(Supplemental file 1; Supplemental Table 2)**.

**Figure 2.**
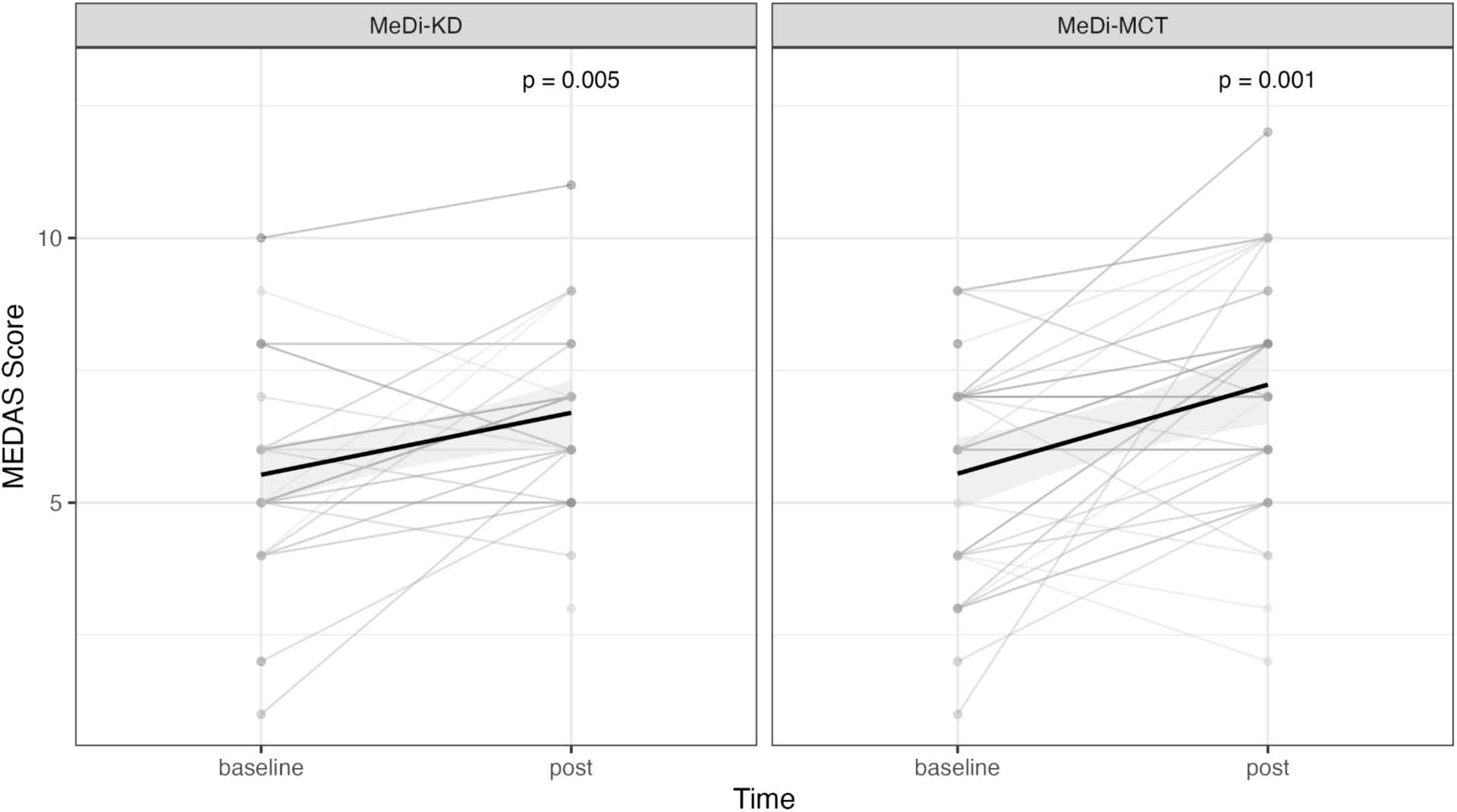
MEDAS scores increase from baseline during each intervention phase. Black lines represent means, gray shading represents the 95% confidence interval, and gray lines represent individual trajectories.

#### Blood BHB

Participant-monitored non-fasting blood BHB concentrations were consistently higher during the MeDi-KD phase compared with the MeDi-MCT phase throughout all 8 weeks of the intervention (p<0.001 **(Figure 3A)**. This significant difference held following removal of outliers (p<0.001), and adjustment for period effects and ketone levels at baseline (p<0.001) in sensitivity analysis. Average non-fasting blood [BHB] in the MeDi-KD group consistently exceeded the threshold of 0.3 mM each week, with a mean of 0.51 (SD: 0.36) mM across study weeks. In contrast, only week 5 and week 7 average [BHB] readings were above 0.3 mM in the MeDi-MCT group, with an overall mean of 0.19 (SD: 0.11) mM. The proportion of participants with mean at-home ketone levels >0.3 mM was 71.4% in the MeDi-KD group, as compared with 22.9% in the MeDi-MCT group (p<0.001).

**Figure 3.**
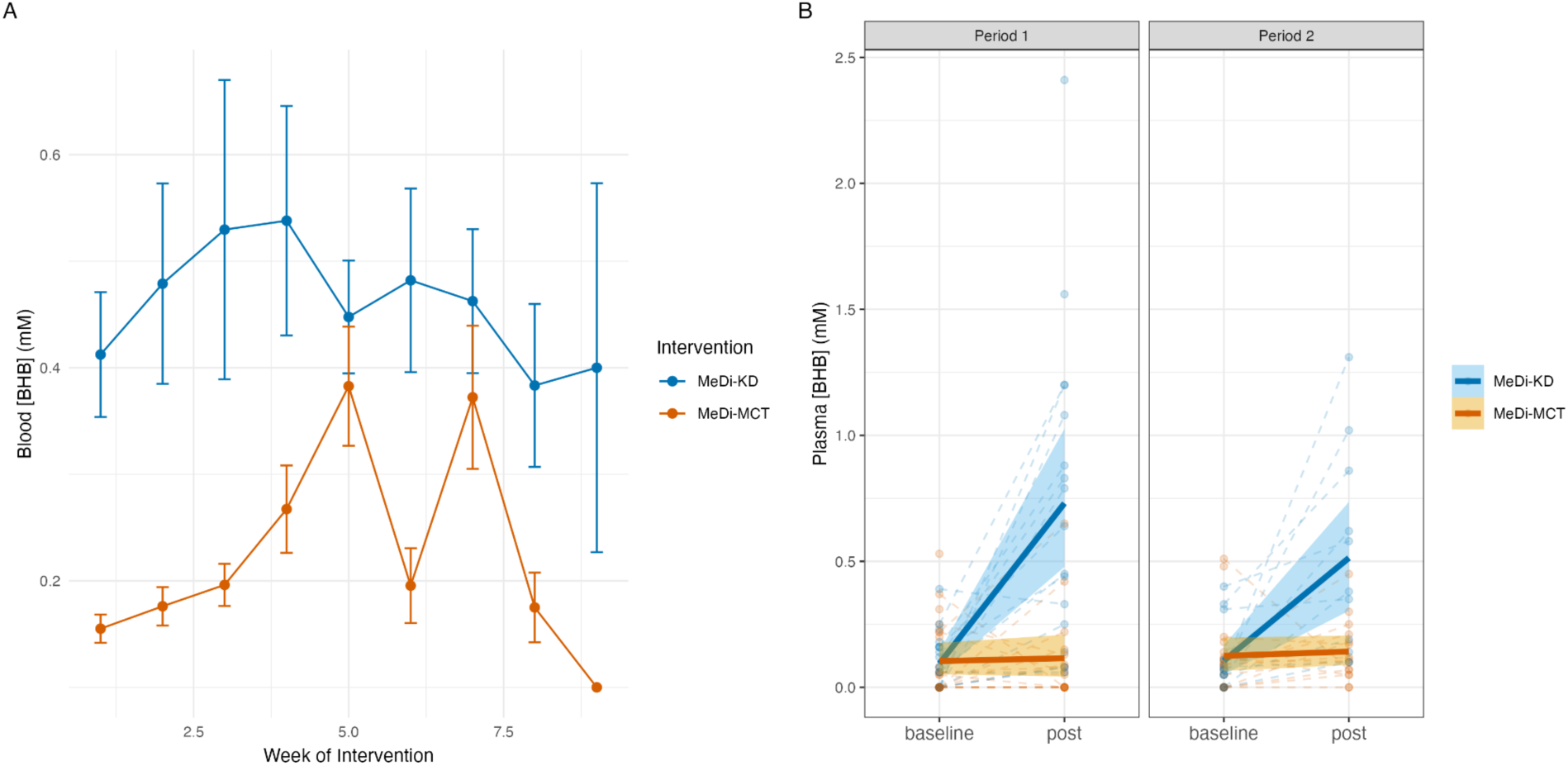
Ketone levels increase relative to baseline following MeDi-KD, but not MeDi-MCT, regimens. **A)** At-home non-fasting blood [BHB] (mM) over the course of each 8-week intervention. Error bars represent standard error of the mean (SEM). **B)** Laboratory measurements of fasting plasma [BHB] (mM) at baseline and post-intervention follow-up visits. Solid lines represent means surrounded by 95% confidence intervals.

Moderate significant correlations between changes from baseline in non-fasting at-home and fasting laboratory ketone measurements were observed for the MeDi-KD (r=0.65; p<0.001) but not the MeDi-MCT (r=0.17; p=0.42) intervention. Fasting plasma [BHB] increased by a mean of 0.45 (SD: 0.41) mM from baseline following the MeDi-KD (p<0.001) to a mean of 0.65 (SD: 0.55) mM at post-intervention visits, with 50% of participants having post-intervention fasting ketone levels >0.5 mM. In contrast, fasting plasma [BHB] rose by only 0.04 (SD: 0.19) mM (p=0.18) following the MeDi-MCT **(Figure 3B)** to a mean of 0.13 (SD: 0.15) mM, with only 1 participant (3%) having a post-intervention fasting ketone level above 0.5 mM. As such, a significant difference was found in the effects of each intervention on fasting plasma [BHB] (p<0.001).

#### Self-Rated Adherence

Among participants completing both intervention phases, 73% reported following the MeDi-KD and 92% reported following the MeDi-MCT almost every day or skipping at most several times per month **(Table 2)**. Self-rated adherence (p=0.2) and likelihood of future adherence (p=0.2) did not differ significantly between MeDi-MCT and MeDi-KD interventions. However, participants took significantly longer to adjust to the MeDi-KD regimen (p=0.006), and rated their likelihood of adhering to a Mediterranean diet alone significantly higher than their likelihood of adhering to a ketogenic (p<0.001) or MCT-supplemented (p<0.001) diet.

**Table 2.** Self-Rated Adherence, Adjustment Period and Likelihood of Future Dietary Adherence.

| Characteristic | MeDi-KD<br>N = 33<sup>1</sup> | MeDi-MCT<br>N = 33<sup>1</sup> |
| --- | --- | --- |
| Self-Rated Adherence |  |  |
| No problem at all, doing it every single day | 12 (46%) | 12 (44%) |
| No real problem but I skip several times a month | 7 (27%) | 13 (48%) |
| Difficult, I skip more than one time a week | 6 (23%) | 2 (7.4%) |
| Very difficult, I can barely execute it | 1 (3.8%) | 0 (0%) |
| Weeks to Adjust to Diet |  |  |
| 0 | 9 (33%) | 21 (88%) |
| 1 | 2 (7.4%) | 1 (4.2%) |
| 2 | 7 (26%) | 2 (8.3%) |
| 3 | 1 (3.7%) | 0 (0%) |
| 4 | 1 (3.7%) | 0 (0%) |
| 6 | 1 (3.7%) | 0 (0%) |
| 8 | 1 (3.7%) | 0 (0%) |
| Never | 5 (19%) | 0 (0%) |
| Likelihood of Future Adherence to Diet |  |  |
| Not likely at all | 6 (21%) | 5 (29%) |
| Unlikely | 4 (14%) | 1 (5.9%) |
| Neutral / Unsure | 14 (50%) | 4 (24%) |
| Likely | 3 (11%) | 6 (35%) |
| Will definitely adhere | 1 (3.6%) | 1 (5.9%) |

<sup>1</sup>n (%)

When ranked into quartiles according to the proportion of non-trace (>0.3 mM) readings, participants’ at-home blood ketone levels were moderately correlated with self-rated adherence to the MeDi-KD (r=0.50; p=0.007), but not the MeDi-MCT (r=-0.09; p=0.66). No significant correlations were observed between MEDAS scores and self-rated adherence to either diet.

### 5. Safety

#### Adverse Events

Eight intervention-related adverse events were reported in the MeDi-KD arm and 10 were reported in the MeDi-MCT arm **(Table 3).** None were classified as serious adverse events. In the MeDi-KD arm, recurrent episodes of hypotension were reported by two participants, in association with vigorous exercise. Intermittent, mild tremor exacerbations were reported by two additional participants on the MeDi-KD, and one participant on the MeDi-MCT. Indigestion was reported by three participants on the MeDi-MCT and one on the MeDi-KD. Diarrhea was reported by two participants on the MeDi-MCT. Other symptoms reported with plausible causal links to the interventions included worsened fatigue and headaches on the MeDi-MCT, insomnia on the MeDi-KD, and a delayed response to levodopa medication on the MeDi-KD. One participant reported an allergic reaction deemed unrelated to the study diet on the MeDi-KD.

**Table 3.** Adverse Events.

| **Diet** | **Adverse Event** | **Severity<sup>1</sup>** | **Pattern<sup>2</sup>** | **Serious Adverse Event (y/n)<sup>3</sup>** | **Causality<sup>4</sup>** |
| --- | --- | --- | --- | --- | --- |
| MeDi-KD | Hypotension | 2 | 2 | N | 3 |
| MeDi-KD | Insomnia | 1 | 2 | N | 2 |
| MeDi-KD | Tremor exacerbation | 1 | 2 | N | 2 |
| MeDi-KD | Tremor exacerbation | 1 | 2 | N | 2 |
| MeDi-KD | Delayed onset of response to levodopa | 2 | 2 | N | 3 |
| MeDi-KD | Indigestion | 1 | 2 | N | 2 |
| MeDi-KD | Allergic Reaction | 1 | 3 | N | 0 |
| MeDi-KD | Hypotension | 1 | 2 | N | 3 |
| MeDi-MCT | Unrelated hospitalization (insomnia & psychotic symptoms post-DBS) | 3 | 1 | Y | 0 |
| MeDi-MCT | Unrelated hospitalization (bike accident) | 3 | 1 | Y | 0 |
| MeDi-MCT | Diarrhea | 1 | 2 | N | 3 |
| MeDi-MCT | Diarrhea | 1-2 | 2 | N | 3 |
| MeDi-MCT | Headache | 1 | 2 | N | 2 |
| MeDi-MCT | Indigestion | 1 | 2 | N | 3 |
| MeDi-MCT | Indigestion | 1 | 2 | N | 3 |
| MeDi-MCT | Tremor exacerbation | 1 | 2 | N | 2 |
| MeDi-MCT | Indigestion | 2-3 | 2 | N | 3 |
| MeDi-MCT | Fatigue | 1 | 3 | N | 1 |

<sup>1</sup>1=Mild; 2=Moderate; 3=Severe.
<sup>2</sup>1=Once; 2=Intermittent; 3=Constant.
<sup>3</sup>Death, hospitalization or incapacity related to the intervention.
<sup>4</sup>0=None; 1=Unlikely; 2=Possible; 3=Probable; 4=Definite.

All participants maintained blood ketone levels within or below the range for physiologic ketosis throughout the study, with the exception of one participant who obtained blood ketone readings of 5.1 mM and 3.1 mM on the same day when following the MeDi-KD, after skipping a midday meal.

#### Fasting Lipids

No significant changes from baseline were observed in fasting triglycerides, LDL, HDL or total cholesterol following either intervention **(Figure 4 A-D).**

**Figure 4.**
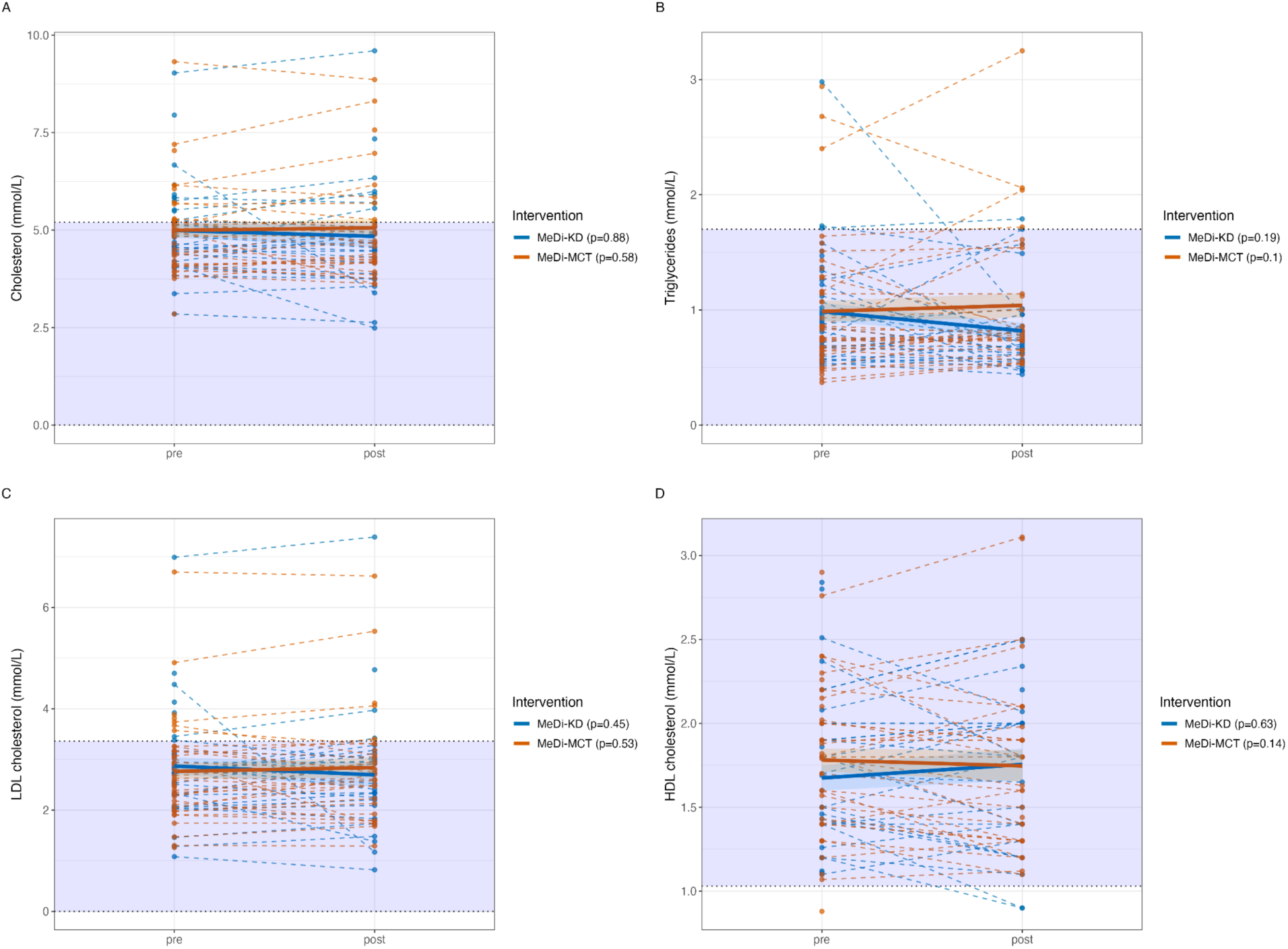
Intervention-associated changes in triglyceride, LDL, HDL and total cholesterol. Purple shading indicates the normal range for each laboratory value.^70^

#### Body Mass

A significant reduction in body mass of a mean -2.63 (SD: 1.86) kg from baseline was observed following the MeDi-KD phase (p<0.001), whereas body mass remained stable during the MeDi-MCT phase (mean change from baseline: 0.03 (SD: 1.74) kg; p=0.61) **(Figure 5)**. In available-case analysis, there was a significant between-intervention difference in weight changes (p<0.001) after adjusting for period effects. Weight loss did not result in an underweight status (BMI <18.5) for any participants in our study at any of the assessments.

**Figure 5.**
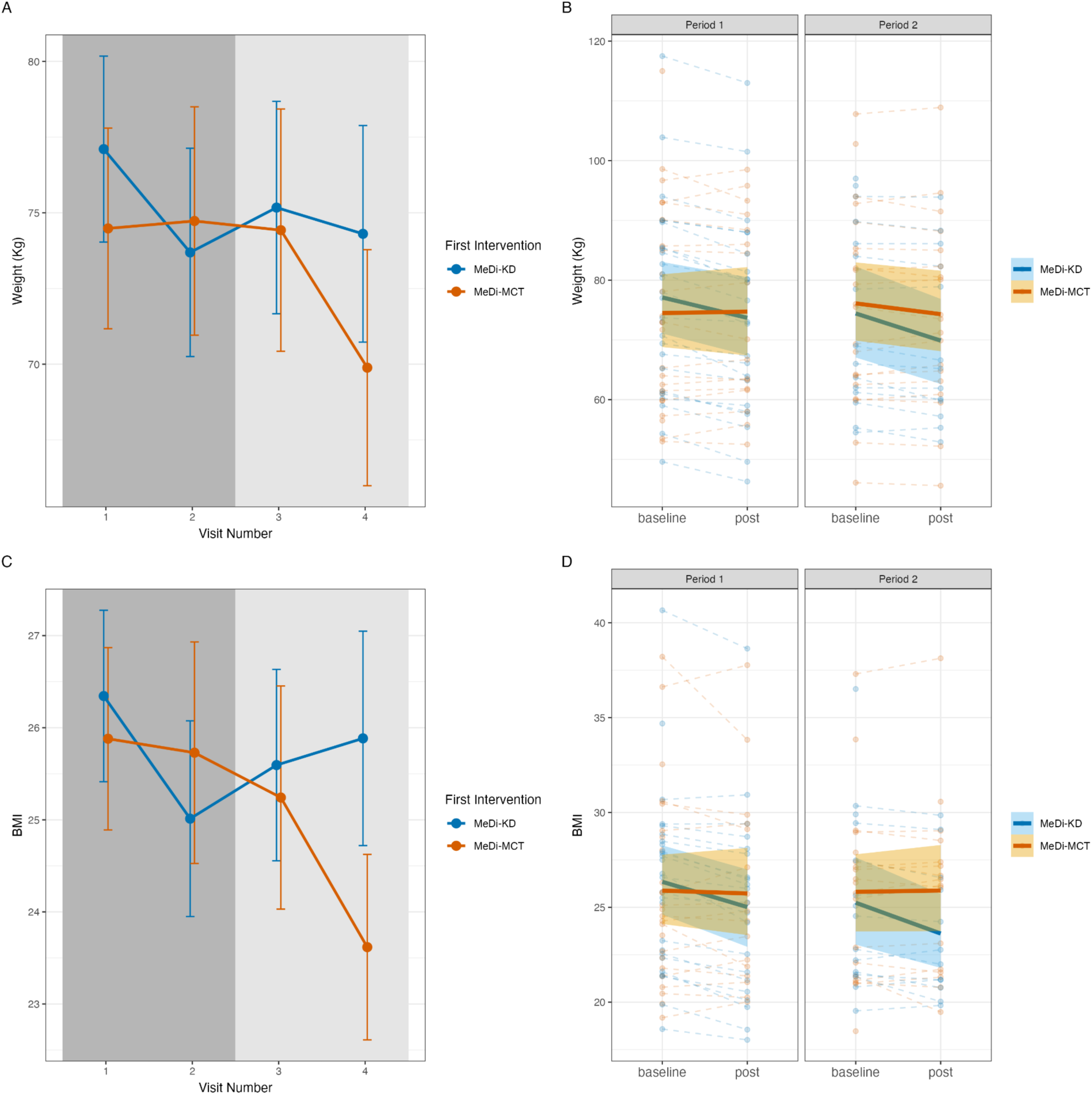
Modest weight loss occurred during the MeDi-KD, but not the MeDi-MCT phase. **A and C)** Mean body mass (A) and BMI (C) over the course of the four study visits. Error bars represent SEM. **B and D)** Changes from baseline in body mass (B) and BMI (D) for each intervention and period. Solid lines represent means surrounded by 95% confidence intervals.

### 6. Clinical Outcomes

A significant mean within-participant decrease in MDS-UPDRS Part II scores of -1.4 (SD: 4.2; p=0.039) points and Part IV scores of -1.0 (SD: 3.0; p=0.044) points was observed following the MeDi-MCT intervention **(Table 4)**. Non-significant improvements in MDS-UPDRS Parts II and IV scores were seen following the MeDi-KD intervention. No between-intervention differences were observed in any parts of the MDS-UPDRS nor in MDS-UPDRS total scores. No between- or within-intervention differences were observed in total MoCA scores. A statistically significant, but clinically negligible,^71^ decrease of two out of a maximum of 134 points on the PD-CRS (p=0.024) was observed following the MeDi-KD in available-case analysis, which was not reproduced in the intention-to-treat analysis. No changes to anti-Parkinsonian regimens were reported between assessments.

**Table 4.**
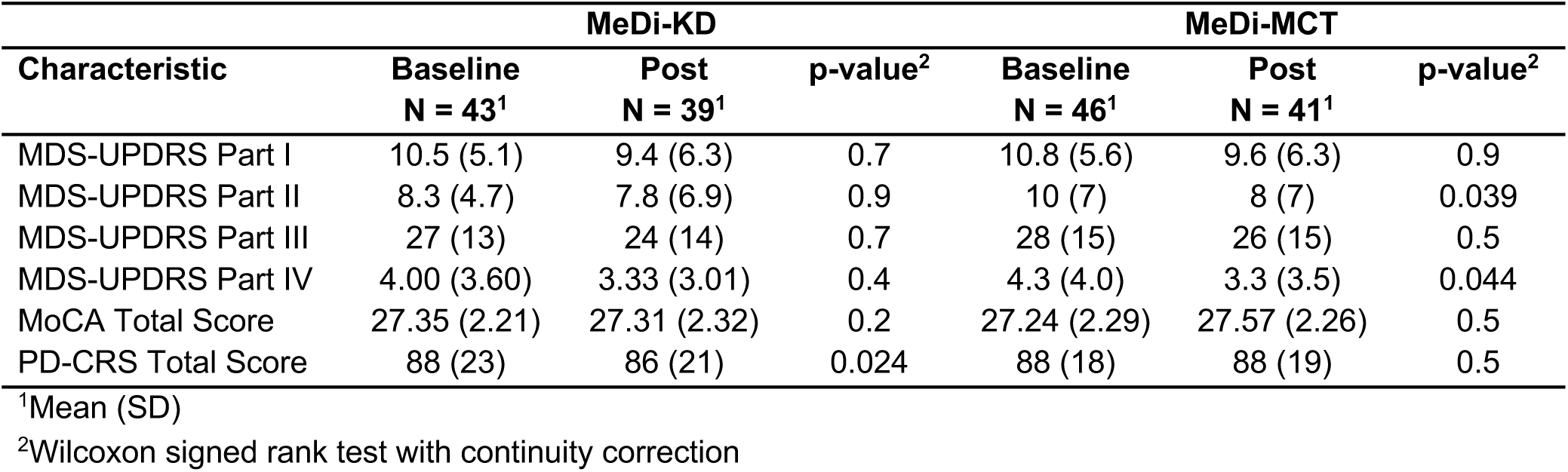
Changes in MDS-UPDRS, MoCA and PD-CRS scores.

| Characteristic | MeDi-KD |  |  | MeDi-MCT |  |  |
| --- | --- | --- | --- | --- | --- | --- |
|  | Baseline<br>N = 43<sup>1</sup> | Post<br>N = 39<sup>1</sup> | p-value<sup>2</sup> | Baseline<br>N = 46<sup>1</sup> | Post<br>N = 41<sup>1</sup> | p-value<sup>2</sup> |
| MDS-UPDRS Part I | 10.5 (5.1) | 9.4 (6.3) | 0.7 | 10.8 (5.6) | 9.6 (6.3) | 0.9 |
| MDS-UPDRS Part II | 8.3 (4.7) | 7.8 (6.9) | 0.9 | 10 (7) | 8 (7) | 0.039 |
| MDS-UPDRS Part III | 27 (13) | 24 (14) | 0.7 | 28 (15) | 26 (15) | 0.5 |
| MDS-UPDRS Part IV | 4.00 (3.60) | 3.33 (3.01) | 0.4 | 4.3 (4.0) | 3.3 (3.5) | 0.044 |
| MoCA Total Score | 27.35 (2.21) | 27.31 (2.32) | 0.2 | 27.24 (2.29) | 27.57 (2.26) | 0.5 |
| PD-CRS Total Score | 88 (23) | 86 (21) | 0.024 | 88 (18) | 88 (19) | 0.5 |

<sup>1</sup>Mean (SD)
<sup>2</sup>Wilcoxon signed rank test with continuity correction

## DISCUSSION

The KIM trial was designed as a response to patient inquiries regarding the utility of ketogenic diets as adjunct management strategies for their PD. This interest presumably stemmed from growing body of pilot data suggesting variable beneficial effects of ketogenic diets on motor, non-motor, neuropsychiatric and cognitive symptoms of PD,^13,15–19^ and our research group’s previous work on diet in PD.^39^ Compelling as is the prospect of a complementary dietary self-management strategy for PD,^8,72^ none of these studies comprehensively assessed the potential long-term risks and barriers to implementation of ketogenic regimens in the PwP. The overarching goal of the KIM trial was to assess the feasibility and safety of both a dietary and MCT supplement-based ketogenic intervention in PwP, optimized from both an overall dietary quality and adherence perspective through their combination with Mediterranean diet principles.^47,73^ We demonstrate the feasibility of recruitment for this 6-month crossover study in an urban setting and the low short-term risk of major adverse health effects of these dietary approaches in PwP. However, we note a higher-than-expected study dropout rate (37% vs. 25%) and constraints on the levels of objective and self-reported markers of adherence achieved with either intervention.

The success of our recruitment effort attests to the high levels of interest in lifestyle self-management strategies among PwP seen at our tertiary Movement Disorders Clinic (Vancouver, British Columbia, Canada), as well as to the overall appropriateness of our eligibility criteria, screening and recruitment procedures.^74–76^ The identification of potentially interested and eligible participants by their neurologists during centre clinic-research hybrid meetings was particularly helpful in prioritizing initial contacts. A notable limitation to our recruitment strategy is reflected in the lack of demographic diversity of our cohort, constituted primarily by Caucasian males with post-secondary education living in proximity to our university campus. Established barriers associated with trial participation in PwP - including language barriers among ethnic minorities, travel distance, financial constraints, work conflicts, and reduced access to caregiver support among women - no doubt impacted our ability to recruit a demographically representative sample.^77^ The apparent lack of cultural adaptability of both the Mediterranean,^78^ and ketogenic diets,^79^ employed in this study may have further contributed to the overrepresentation of those accustomed to a Western-style eating pattern in our cohort.

Similar participant and intervention-specific factors may have contributed to our sub-target retention rate. Compliance to standard ketogenic diet regimens is notoriously challenging. Even in the case of intractable epilepsy, which represents the strongest current indication for the ketogenic diet in adults, retention rates at 6 months in intervention studies are reported as low as 38-46%.^80–83^ In one study, over one-third of patients with no alternative treatment options for refractory epilepsy nevertheless refused to participate in a ketogenic diet intervention due to its perceived restrictiveness and complexity.^83^ In PD, reported completion rates for 8-12 week ketogenic diet trials ranges from 70-88%, with ours being the first ketogenic intervention study longer than 3 months undertaken in this population.

While some evidence has indicated higher retention rates to Mediterranean adaptations of the ketogenic diet in adults with overweight, obesity and MCI,^47,84,85^ our retention data indicates unresolved challenges to its adoption by PwP. Particularly important to consider in PwP are issues related to the effects of high fat intakes on gastric motility and dopaminergic medication absorption, as was illustrated by one withdrawal from the MeDi-KD arm attributed to a delay in levodopa onset of action. Gastrointestinal disturbances may help to account for our lack of observed advantage of the MeDi-MCT over the MeDi-KD in minimizing dropouts, whereas retention rates as high as 95% have been reported for long-term MCT supplementation trials in older adults with probable Alzheimer’s disease.^86^ Specifically, the high rates of baseline gastrointestinal dysfunction in PD may make a subset of PwP particularly susceptible to more severe adverse gastrointestinal side effects of MCT oil.^86,87^ In our study, two out of a total of five such adverse events were sufficiently severe to prompt withdrawal from the MeDi-MCT arm.

Apart from gastrointestinal side effects, other adverse events deemed possibly or probably related to the study diets included hypotension (MeDi-KD) and tremor exacerbations (both MeDi-KD and MeDi-MCT). Hypotension is a known potential adverse effect related to KD-induced diuresis,^29^ both of which may be of particular concern in PwP who often experience urinary dysfunction,^88^ orthostatic hypotension,^89^ and are at elevated risk of falls.^90^ Clear instruction regarding fluid intake is therefore essential for any PwP interested in following a KD.^17,91^ Transient KD-related tremor exacerbations have been previously documented in PwP^17^; however, the underlying mechanism remains unclear. From a cardiovascular risk perspective, the emphasis on unsaturated fats in the MeDi-KD appears to have mitigated against short-term KD-associated spikes in total and LDL cholesterol in our cohort on aggregate.^59^ However, previously reported beneficial effects of Mediterranean-ketogenic diets on HDL-C were not observed in our study.^50,84,92^ Notably, there was considerable inter-individual heterogeneity in lipid profile changes induced by both diets, emphasizing the need for close monitoring of blood lipid levels in future ketogenic diet trials.^59^ The modest weight loss observed in the MeDi-KD arm did not place any participants in a higher risk category according to BMI status in our study; however, unintended weight loss may make the ketogenic diet a poor option for PwP struggling to maintain adequate body mass.^93,94^ Together, our findings suggest that combining ketogenic and Mediterranean principles minimize some but not all of the safety risks associated with standard KDs.

The apparent improved safety profile of the MeDi-KD relative to standard KDs may come at the expense of “optimal” levels of nutritional ketosis (1.5-3.0 mM) achievable with more severe carbohydrate restriction.^73,95^ Nevertheless, at-home and laboratory based blood BHB measurements demonstrated that the majority of participants in our study were able to sustain “light” nutritional ketosis (0.3-1.0 mM post-prandial; 0.5-1.0 mM fasting) while following the MeDi-KD, comparable to levels observed in previous KD studies in PwP.^16,96^ Importantly, expected thresholds for nutritional ketosis with KDs of varying macronutrient composition have yet to be established in PwP.^16^ Disease-specific thresholds may be relevant, as previous studies have demonstrated substantial inter-individual variability in responses to diets of identical ketogenic ratios, with older age representing a constraint on the levels of ketosis achievable.^97^ Explanations for this phenomenon may relate to age-dependent elevations in postprandial insulin, decreases in plasma carnitine, decreases in plasma lipoprotein lipase activity and decreases in hepatic glucagon sensitivity.^98^

High inter-individual variability in the ketogenic effect of MCT oil has likewise been demonstrated in human studies.^98^ MCT supplementation is known to induce at most mild ketonemia in the absence of dietary carbohydrate and/or caloric restriction,^12,99–101^ with several studies reporting no MCT-induced increases in plasma,^102^ or brain BHB.^101,103^ The subthreshold blood BHB elevations seen in our MeDi-MCT intervention, and their dissociation from self-rated adherence scores, was therefore not entirely unexpected, and were not interpreted as evidence that the MCT oil was not consumed as reported. Importantly, blood [BHB] may also be a poor predictor of the likelihood of neurologic benefit from MCT supplementation: emerging evidence suggests that MCTs may exert neuroprotective effects via the direct actions of plasma and brain medium-chain fatty acids,^12,101^ independent of changes in circulating ketones. Together, these results point to the need for future studies of MCT supplementation to assess alternative objective adherence measures to blood ketones, such as plasma medium chain fatty acids, whose levels may better correlate with primary bioenergetic and/or clinical outcomes.

Interestingly, self-rated adherence was also not associated with MEDAS scores during either intervention. Despite participants overwhelmingly claiming to have deviated from each diet program no more than a few times per week, MEDAS scores were predominantly in the “moderate adherence” category. Past studies have demonstrated similar disconnects between objective and subjective measures of adherence to Mediterranean diets.^104^ Adherence may be remediable with more intensive dietary coaching, involving care partners where possible,^105^ supplying ingredients or meal delivery, or by including a run-in period to test adherence and then only including those with satisfactory adherence in the main trial.^66,73,85,106^ The apparent ceiling effect for changes in MEDAS scores on the MeDi-KD may be additionally explained by the combined diet’s inherent restriction of carbohydrates like fruits, vegetables, and legumes, which figure in to the MEDAS calculation.

### Exploratory Efficacy

Our secondary analyses revealed significant improvements in MDS-UPDRS Parts II and IV scores following the MeDi-MCT intervention, with similar trends observed in the MeDi-KD phase. Decreases in MDS-UPDRS Parts II and IV scores following a ketogenic diet was reported in a previous RCT in PwP.^17^ While the mechanisms underlying these observations are not fully understood, improved gastrointestinal motility reported by several participants in our study may have optimized levodopa pharmacokinetics, alleviating motor complications and improving motor experience of daily living.^107^ Diet-related changes in gut microbiota composition and metabolic function may have also contributed to this effect.^17,107^ Analysis of diet-induced shifts in the gut microbiota during the KIM trial is currently underway, and will be reported in a future publication. Finally, the unblinded nature of the trial makes it essential to consider the influence of participant expectations on clinical outcomes, particularly given that changes were observed only in self-reported parts of the MDS-UPDRS, and not in the objectively-rated MDS-UPDRS Part III.^108^ The absence of any clinically meaningful changes in cognition scores was unsurprising given the short interval between baseline and follow-up visits, and the underrepresentation of participants with baseline cognitive impairment in our study.^109^

### Limitations

Several limitations need to be considered in evaluating our study findings. While we did not observe adverse effects of either diet on fasting lipid profiles nor major adverse events, our small sample size and short-duration interventions lower the likelihood that our study would have been able to detect such effects, as well as rarer or long-term complications of ketogenic diets and/or MCT supplementation, including vitamin and mineral deficiencies, osteopaenia and kidney stones.^21^ Future, longer-duration studies involving more precise assessment of dietary intake would be better poised to assess the true safety impacts of sustained nutritional ketosis in PwP. Likewise, such studies would help to clarify whether the expected levels of ketosis for given macronutrient compositions differ in PwP relative to the general population.

## Conclusion

Mediterranean-ketogenic and MCT-supplemented Mediterranean diets appear to be generally safe in individuals with mild-to-moderate PD. Mild nutritional ketosis was achieved by approximately one-half of participants following the carbohydrate-restricted MeDi-KD, but was not observed with MCT supplementation in the absence of carbohydrate restriction during the MeDi-MCT. Modest observed improvements in motor experiences of daily living and motor complications warrant validation in phase III studies. Gastrointestinal side effects and cholesterol levels should be monitored closely in any such future trials, or in clinical practice for any patients opting to follow a ketogenic or MCT-supplemented diet.

## ACKNOWLEDGEMENTS

The authors would like to thank all personnel who supported the trial’s operations, and all the participants for the time and effort invested in our study. Dr. Biljana Jonoska Stojkova and Frances Cheng are thanked for their statistical support. Dr. Christopher Gardner and Dalia Perelman of Stanford Medicine are thanked for their guidance around the development of participant diet education materials. We would additionally like to acknowledge Ajinomoto Cambrooke Inc. (United States), a therapeutic nutrition company, which supplied the gram scales for participants and offered two Ketogenic Diet Professional Training sessions to YNW.

## AUTHOR CONTRIBUTIONS

SK, MSC, AMR, MAS, BBF, TRC and SAC were involved in study conceptualization, methodology and funding acquisition. KT, YNW and AJK were responsible for investigation, data curation and project administration. KT conducted the formal analysis. KT, JSTL and YNW prepared the original draft of the manuscript. All authors reviewed and approved of the final version of the manuscript. SAC supervised the project.

## STATEMENTS AND DECLARATIONS

### Ethical considerations

Ethical approval was granted by the University of British Columbia Clinical Research Ethics Board (Protocol #: H21-03747).

### Consent to participate

All participants in the study provided written informed consent prior to enrolment.

### Consent for publication

All participants consented to the de-identified dissemination of study findings as a part of the informed consent process.

### Data availability

The data supporting the findings of this study are available upon reasonable request to the corresponding author.

## SUPPLEMENTAL MATERIAL

The supplemental material is available in the electronic version of this article.

## REFERENCES

1. Luo Y, Qiao L, Li M, et al. Global, regional, national epidemiology and trends of Parkinson’s disease from 1990 to 2021: findings from the Global Burden of Disease Study 2021. Front Aging Neurosci 2024; 16: 1498756.

2. Dorsey ER, Constantinescu R, Thompson JP, et al. Projected number of people with Parkinson disease in the most populous nations, 2005 through 2030. Neurology 2007; 68: 384–386.

3. Dorsey ER, Sherer T, Okun MS, et al. The emerging evidence of the Parkinson pandemic. J Parkinsons Dis 2018; 8: S3–S8.

4. Su D, Cui Y, He C, et al. Projections for prevalence of Parkinson’s disease and its driving factors in 195 countries and territories to 2050: modelling study of Global Burden of Disease Study 2021. BMJ 2025; e080952.

5. Vijiaratnam N, Simuni T, Bandmann O, et al. Progress towards therapies for disease modification in Parkinson’s disease. Lancet Neurol 2021; 20: 559–572.

6. van der Kolk NM, de Vries NM, Kessels RPC, et al. Effectiveness of home-based and remotely supervised aerobic exercise in Parkinson’s disease: a double-blind, randomised controlled trial. Lancet Neurol 2019; 18: 998–1008.

7. Choi A, Hallett M, Ehrlich D. Nutritional ketosis in Parkinson’s disease - a review of remaining questions and insights. Neurotherapeutics 2021; 18: 1637–1649.

8. Tosefsky KN, Zhu J, Wang YN, et al. The role of diet in Parkinson’s disease. J Parkinsons Dis 2024; 14: S21–S34.

9. Gough SM, Casella A, Ortega KJ, et al. Neuroprotection by the ketogenic diet: Evidence and controversies. Front Nutr 2021; 8: 782657.

10. Kim J-M. Ketogenic diet: Old treatment, new beginning. Clin Neurophysiol Pract 2017; 2: 161–162.

11. St-Pierre V, Vandenberghe C, Lowry C-M, et al. Plasma ketone and medium chain fatty acid response in humans consuming different medium chain triglycerides during a metabolic study day. Front Nutr 2019; 6: 46.

12. Courchesne-Loyer A, Fortier M, Tremblay-Mercier J, et al. Stimulation of mild, sustained ketonemia by medium-chain triacylglycerols in healthy humans: estimated potential contribution to brain energy metabolism. Nutrition 2013; 29: 635–640.

13. Koyuncu H, Fidan V, Toktas H, et al. Effect of ketogenic diet versus regular diet on voice quality of patients with Parkinson’s disease. Acta Neurol Belg 2021; 121: 1729–1732.

14. Norwitz NG, Dearlove DJ, Lu M, et al. A ketone ester drink enhances endurance exercise performance in Parkinson’s disease. Front Neurosci 2020; 14: 584130.

15. Tidman MM, White D, White T. Effects of an low carbohydrate/healthy fat/ketogenic diet on biomarkers of health and symptoms, anxiety and depression in Parkinson’s disease: a pilot study. Neurodegener Dis Manag 2022; 12: 57–66.

16. Choi AH, Delgado M, Chen KY, et al. A randomized feasibility trial of medium chain triglyceride-supplemented ketogenic diet in people with Parkinson’s disease. BMC Neurol 2024; 24: 106.

17. Phillips MCL, Murtagh DKJ, Gilbertson LJ, et al. Low-fat versus ketogenic diet in Parkinson’s disease: A pilot randomized controlled trial: Low-Fat Versus Ketogenic Diet in PD. Mov Disord 2018; 33: 1306–1314.

18. Vanitallie TB, Nonas C, Di Rocco A, et al. Treatment of Parkinson disease with diet-induced hyperketonemia: a feasibility study. Neurology 2005; 64: 728–730.

19. Krikorian R, Shidler MD, Summer SS, et al. Nutritional ketosis for mild cognitive impairment in Parkinson’s disease: A controlled pilot trial. Clin Park Relat Disord 2019; 1: 41–47.

20. Kenig S, Petelin A, Poklar Vatovec T, et al. Assessment of micronutrients in a 12-wk ketogenic diet in obese adults. Nutrition 2019; 67–68: 110522.

21. Crosby L, Davis B, Joshi S, et al. Ketogenic diets and chronic disease: Weighing the benefits against the risks. Front Nutr 2021; 8: 702802.

22. Gardner CD, Vadiveloo MK, Petersen KS, et al. Popular dietary patterns: Alignment with American Heart Association 2021 dietary guidance: A scientific statement from the American Heart Association. Circulation 2023; 147: 1715–1730.

23. Fereshtehnejad S-M, Ghazi L, Sadeghi M, et al. Prevalence of malnutrition in patients with Parkinson’s disease: a comparative study with healthy controls using Mini Nutritional Assessment (MNA) questionnaire. J Parkinsons Dis 2014; 4: 473–481.

24. Bueno NB, de Melo ISV, de Oliveira SL, et al. Very-low-carbohydrate ketogenic diet v. low-fat diet for long-term weight loss: a meta-analysis of randomised controlled trials. Br J Nutr 2013; 110: 1178–1187.

25. Mansoor N, Vinknes KJ, Veierød MB, et al. Effects of low-carbohydrate diets v. low-fat diets on body weight and cardiovascular risk factors: a meta-analysis of randomised controlled trials. Br J Nutr 2016; 115: 466–479.

26. Gardner CD, Landry MJ, Perelman D, et al. Effect of a ketogenic diet versus Mediterranean diet on glycated hemoglobin in individuals with prediabetes and type 2 diabetes mellitus: The interventional Keto-Med randomized crossover trial. Am J Clin Nutr 2022; 116: 640–652.

27. Kirkpatrick CF, Bolick JP, Kris-Etherton PM, et al. Review of current evidence and clinical recommendations on the effects of low-carbohydrate and very-low-carbohydrate (including ketogenic) diets for the management of body weight and other cardiometabolic risk factors: A scientific statement from the National Lipid Association Nutrition and Lifestyle Task Force. J Clin Lipidol 2019; 13: 689–711.e1.

28. Iatan I, Huang K, Vikulova D, et al. Association of a low-carbohydrate high-fat diet with plasma lipid levels and cardiovascular risk. JACC Adv 2024; 3: 100924.

29. Watanabe M, Tuccinardi D, Ernesti I, et al. Scientific evidence underlying contraindications to the ketogenic diet: An update. Obes Rev 2020; 21: e13053.

30. Akbar U, He Y, Dai Y, et al. Weight loss and impact on quality of life in Parkinson’s disease. PLoS One 2015; 10: e0124541.

31. Rew L, Harris MD, Goldie J. The ketogenic diet: its impact on human gut microbiota and potential consequent health outcomes: a systematic literature review. Gastroenterol Hepatol Bed Bench 2022; 15: 326–342.

32. Romano S, Savva GM, Bedarf JR, et al. Meta-analysis of the Parkinson’s disease gut microbiome suggests alterations linked to intestinal inflammation. NPJ Parkinsons Dis 2021; 7: 27.

33. Wallen ZD, Demirkan A, Twa G, et al. Metagenomics of Parkinson’s disease implicates the gut microbiome in multiple disease mechanisms. Nat Commun 2022; 13: 6958.

34. Toh TS, Chong CW, Lim S-Y, et al. Gut microbiome in Parkinson’s disease: New insights from meta-analysis. Parkinsonism Relat Disord 2022; 94: 1–9.

35. Scheperjans F, Aho V, Pereira PAB, et al. Gut microbiota are related to Parkinson’s disease and clinical phenotype. Mov Disord 2015; 30: 350–358.

36. Taylor MK, Swerdlow RH, Burns JM, et al. An experimental ketogenic diet for Alzheimer disease was nutritionally dense and rich in vegetables and avocado. Curr Dev Nutr 2019; 3: nzz003.

37. Ellouze I, Sheffler J, Nagpal R, et al. Dietary patterns and Alzheimer’s disease: An updated review linking nutrition to neuroscience. Nutrients 2023; 15: 3204.

38. Alcalay RN, Gu Y, Mejia-Santana H, et al. The association between Mediterranean diet adherence and Parkinson’s disease: Association of MeDi Adherence and PD. Mov Disord 2012; 27: 771–774.

39. Metcalfe-Roach A, Yu AC, Golz E, et al. MIND and Mediterranean diets associated with later onset of Parkinson’s disease. Mov Disord 2021; 36: 977–984.

40. Maraki MI, Yannakoulia M, Stamelou M, et al. Mediterranean diet adherence is related to reduced probability of prodromal Parkinson’s disease. Mov Disord 2019; 34: 48–57.

41. Paknahad Z, Sheklabadi E, Derakhshan Y, et al. The effect of the Mediterranean diet on cognitive function in patients with Parkinson’s disease: A randomized clinical controlled trial. Complement Ther Med 2020; 50: 102366.

42. Seelarbokus BA, Menozzi E, Schapira AHV, et al. Mediterranean diet adherence, gut Microbiota and Parkinson’s disease: A systematic review. Nutrients 2024; 16: 2181.

43. Paknahad Z, Sheklabadi E, Moravejolahkami AR, et al. The effects of Mediterranean diet on severity of disease and serum Total Antioxidant Capacity (TAC) in patients with Parkinson’s disease: a single center, randomized controlled trial. Nutr Neurosci 2022; 25: 313–320.

44. Fox DJ, Park SJ, Mischley LK. Comparison of associations between MIND and Mediterranean diet scores with patient-reported outcomes in Parkinson’s disease. Nutrients 2022; 14: 5185.

45. Rusch C, Beke M, Tucciarone L, et al. Mediterranean diet adherence in people with Parkinson’s disease reduces constipation symptoms and changes fecal Microbiota after a 5-week single-arm pilot study. Front Neurol 2021; 12: 794640.

46. Rusch C, Beke M, Nieves C Jr, et al. Promotion of a Mediterranean diet alters constipation symptoms and fecal calprotectin in people with Parkinson’s disease: A randomized controlled trial. Nutrients 2024; 16: 2946.

47. Nagpal R, Neth BJ, Wang S, et al. Modified Mediterranean-ketogenic diet modulates gut microbiome and short-chain fatty acids in association with Alzheimer’s disease markers in subjects with mild cognitive impairment. EBioMedicine 2019; 47: 529–542.

48. Kumar A, Sharma M, Su Y, et al. Small extracellular vesicles in plasma reveal molecular effects of modified Mediterranean-ketogenic diet in participants with mild cognitive impairment. Brain Commun 2022; 4: fcac262.

49. Neth BJ, Huynh K, Giles C, et al. Consuming a modified Mediterranean ketogenic diet reverses the peripheral lipid signature of Alzheimer’s disease in humans. Commun Med (Lond) 2025; 5: 11.

50. Schweickart A, Batra R, Neth BJ, et al. Serum and CSF metabolomics analysis shows Mediterranean Ketogenic Diet mitigates risk factors of Alzheimer’s disease. NPJ Metab Health Dis 2024; 2: 15.

51. Phillips MCL, Deprez LM, Mortimer GMN, et al. Randomized crossover trial of a modified ketogenic diet in Alzheimer’s disease. Alzheimers Res Ther 2021; 13: 51.

52. Taylor MK, Sullivan DK, Mahnken JD, et al. Feasibility and efficacy data from a ketogenic diet intervention in Alzheimer’s disease. Alzheimers Dement (N Y) 2018; 4: 28–36.

53. Martin-McGill KJ, Bresnahan R, Levy RG, et al. Ketogenic diets for drug-resistant epilepsy. Cochrane Database Syst Rev 2020; 6: CD001903.

54. Li S, Du Y, Meireles C, et al. Adherence to ketogenic diet in lifestyle interventions in adults with overweight or obesity and type 2 diabetes: a scoping review. Nutr Diabetes 2023; 13: 16.

55. Carrera-Juliá S, Estrela JM, Zacarés M, et al. Effect of the Mediterranean diet supplemented with nicotinamide riboside and pterostilbene and/or coconut oil on anthropometric variables in amyotrophic lateral sclerosis. A pilot study. Front Nutr 2023; 10: 1232184.

56. Goetz CG, Fahn S, Martinez-Martin P, et al. Movement Disorder Society-sponsored revision of the Unified Parkinson’s Disease Rating Scale (MDS-UPDRS): Process, format, and clinimetric testing plan. Mov Disord 2007; 22: 41–47.

57. Nasreddine ZS, Phillips NA, Bédirian V, et al. The Montreal Cognitive Assessment, MoCA: a brief screening tool for mild cognitive impairment: Moca: A brief screening tool for MCI. J Am Geriatr Soc 2005; 53: 695–699.

58. Pagonabarraga J, Kulisevsky J, Llebaria G, et al. Parkinson’s disease-cognitive rating scale: a new cognitive scale specific for Parkinson’s disease: Cognitive Rating Scale for PD. Mov Disord 2008; 23: 998–1005.

59. Popiolek-Kalisz J. Ketogenic diet and cardiovascular risk - state of the art review. Curr Probl Cardiol 2024; 49: 102402.

60. Leow ZZX, Guelfi KJ, Davis EA, et al. The glycaemic benefits of a very-low-carbohydrate ketogenic diet in adults with Type 1 diabetes mellitus may be opposed by increased hypoglycaemia risk and dyslipidaemia. Diabet Med. Epub ahead of print 8 May 2018. DOI: 10.1111/dme.13663.

61. Human Microbiome Project Consortium. A framework for human microbiome research. Nature 2012; 486: 215–221.

62. Csizmadi I, Boucher BA, Lo Siou G, et al. Using national dietary intake data to evaluate and adapt the US Diet History Questionnaire: the stepwise tailoring of an FFQ for Canadian use. Public Health Nutr 2016; 19: 3247–3255.

63. Harris JA, Benedict FG. A biometric study of human basal metabolism. Proc Natl Acad Sci U S A 1918; 4: 370–373.

64. Schröder H, Fitó M, Estruch R, et al. A short screener is valid for assessing Mediterranean diet adherence among older Spanish men and women. J Nutr 2011; 141: 1140–1145.

65. Gibson AA, Eroglu EI, Rooney K, et al. Urine dipsticks are not accurate for detecting mild ketosis during a severely energy restricted diet. Obes Sci Pract 2020; 6: 544–551.

66. Sheffler JL, Arjmandi B, Quinn J, et al. Feasibility of an MI-CBT ketogenic adherence program for older adults with mild cognitive impairment. Pilot Feasibility Stud 2022; 8: 16.

67. Miller LE, Ibarra A, Ouwehand AC, et al. Normative values for stool frequency and form using Rome III diagnostic criteria for functional constipation in adults: systematic review with meta-analysis. Ann Gastroenterol 2017; 30: 161–167.

68. Wong RK, Palsson OS, Turner MJ, et al. Inability of the Rome III criteria to distinguish functional constipation from constipation-subtype irritable bowel syndrome. Am J Gastroenterol 2010; 105: 2228–2234.

69. García-Conesa M-T, Philippou E, Pafilas C, et al. Exploring the validity of the 14-item Mediterranean Diet Adherence Screener (MEDAS): A cross-national study in seven European countries around the Mediterranean region. Nutrients 2020; 12: 2960.

70. White-Al Habeeb NMA, Higgins V, Venner AA, et al. Canadian Society of Clinical Chemists harmonized clinical laboratory lipid reporting recommendations on the basis of the 2021 Canadian Cardiovascular Society lipid guidelines. Can J Cardiol 2022; 38: 1180–1188.

71. Fernández de Bobadilla R, Pagonabarraga J, Martínez-Horta S, et al. Parkinson’s disease-cognitive rating scale: psychometrics for mild cognitive impairment: PD-CRS Responsiveness And Cutoffs For PD-MCI. Mov Disord 2013; 28: 1376–1383.

72. van der Berg I, Schootemeijer S, Overbeek K, et al. Dietary interventions in Parkinson’s disease. J Parkinsons Dis 2024; 14: 1–16.

73. Landry MJ, Crimarco A, Perelman D, et al. Adherence to ketogenic and Mediterranean study diets in a crossover trial: The Keto-Med randomized trial. Nutrients 2021; 13: 967.

74. Idnay B, Fang Y, Butler A, et al. Uncovering key clinical trial features influencing recruitment. J Clin Transl Sci 2023; 7: e199.

75. Zahren C, Harvey S, Weekes L, et al. Clinical trials site recruitment optimisation: Guidance from Clinical Trials: Impact and Quality. Clin Trials 2021; 18: 594–605.

76. Newington L, Metcalfe A. Factors influencing recruitment to research: qualitative study of the experiences and perceptions of research teams. BMC Med Res Methodol 2014; 14: 10.

77. Vaswani PA, Tropea TF, Dahodwala N. Overcoming barriers to Parkinson disease trial participation: Increasing diversity and novel designs for recruitment and retention. Neurotherapeutics 2020; 17: 1724–1735.

78. Woodside J, Young IS, McKinley MC. Culturally adapting the Mediterranean Diet pattern - a way of promoting more ‘sustainable’ dietary change? Br J Nutr 2022; 128: 693–703.

79. Seo JH, Kim HD. Cultural challenges in using the ketogenic diet in Asian countries. Epilepsia 2008; 49 Suppl 8: 50–52.

80. Payne NE, Cross JH, Sander JW, et al. The ketogenic and related diets in adolescents and adults--a review: Ketogenic and Related Diets in Adolescents and Adults. Epilepsia 2011; 52: 1941–1948.

81. Ye F, Li X-J, Jiang W-L, et al. Efficacy of and patient compliance with a ketogenic diet in adults with intractable epilepsy: a meta-analysis. J Clin Neurol 2015; 11: 26–31.

82. Green SF, Nguyen P, Kaalund-Hansen K, et al. Effectiveness, retention, and safety of modified ketogenic diet in adults with epilepsy at a tertiary-care centre in the UK. J Neurol 2020; 267: 1171–1178.

83. Mosek A, Natour H, Neufeld MY, et al. Ketogenic diet treatment in adults with refractory epilepsy: a prospective pilot study. Seizure 2009; 18: 30–33.

84. Pérez-Guisado J, Muñoz-Serrano A, Alonso-Moraga A. Spanish Ketogenic Mediterranean Diet: a healthy cardiovascular diet for weight loss. Nutr J 2008; 7: 30.

85. Sheffler JL, Kiosses DN, He Z, et al. Improving adherence to a Mediterranean ketogenic nutrition program for high-risk older adults: A pilot randomized trial. Nutrients 2023; 15: 2329.

86. Juby AG, Blackburn TE, Mager DR. Use of medium chain triglyceride (MCT) oil in subjects with Alzheimer’s disease: A randomized, double-blind, placebo-controlled, crossover study, with an open-label extension. Alzheimers Dement (N Y) 2022; 8: e12259.

87. Warnecke T, Schäfer K-H, Claus I, et al. Gastrointestinal involvement in Parkinson’s disease: pathophysiology, diagnosis, and management. NPJ Parkinsons Dis 2022; 8: 31.

88. Blackett H, Walker R, Wood B. Urinary dysfunction in Parkinson’s disease: a review. Parkinsonism Relat Disord 2009; 15: 81–87.

89. Palma J-A, Kaufmann H. Orthostatic hypotension in Parkinson disease. Clin Geriatr Med 2020; 36: 53–67.

90. Silva de Lima AL, Smits T, Darweesh SKL, et al. Home-based monitoring of falls using wearable sensors in Parkinson’s disease. Mov Disord 2020; 35: 109–115.

91. Hersant H, Grossberg G. The ketogenic diet and Alzheimer’s disease. J Nutr Health Aging 2022; 26: 606–614.

92. Paoli A, Cenci L, Grimaldi KA. Effect of ketogenic Mediterranean diet with phytoextracts and low carbohydrates/high-protein meals on weight, cardiovascular risk factors, body composition and diet compliance in Italian council employees. Nutr J 2011; 10: 112.

93. Ma K, Xiong N, Shen Y, et al. Weight loss and malnutrition in patients with Parkinson’s disease: Current knowledge and future prospects. Front Aging Neurosci 2018; 10: 1.

94. Cersosimo MG, Raina GB, Pellene LA, et al. Weight loss in Parkinson’s disease: The relationship with motor symptoms and disease progression. Biomed Res Int 2018; 2018: 9642524.

95. Phinney SD, Volek JS. The art and science of low carbohydrate living. Beyond Obesity, 2011.

96. Tidman MM, White DR, White TA. Impact of a keto diet on symptoms of Parkinson’s disease, biomarkers, depression, anxiety and quality of life: a longitudinal study. Neurodegener Dis Manag 2024; 14: 97–110.

97. Porper K, Zach L, Shpatz Y, et al. Dietary-induced ketogenesis: Adults are not children. Nutrients 2021; 13: 3093.

98. Juby AG, Brocks DR, Jay DA, et al. Assessing the impact of factors that influence the ketogenic response to varying doses of medium chain triglyceride (MCT) oil. J Prev Alzheimers Dis 2021; 8: 19–28.

99. Henderson ST, Vogel JL, Barr LJ, et al. Study of the ketogenic agent AC-1202 in mild to moderate Alzheimer’s disease: a randomized, double-blind, placebo-controlled, multicenter trial. Nutr Metab (Lond) 2009; 6: 31.

100. Reger MA, Henderson ST, Hale C, et al. Effects of beta-hydroxybutyrate on cognition in memory-impaired adults. Neurobiol Aging 2004; 25: 311–314.

101. Shcherbakova K, Schwarz A, Apryatin S, et al. Supplementation of regular diet with medium-chain triglycerides for procognitive effects: A narrative review. Front Nutr 2022; 9: 934497.

102. Tan KN, Carrasco-Pozo C, McDonald TS, et al. Tridecanoin is anticonvulsant, antioxidant, and improves mitochondrial function. J Cereb Blood Flow Metab 2017; 37: 2035–2048.

103. Wang D, Mitchell ES. Cognition and synaptic-plasticity related changes in aged rats supplemented with 8- and 10-carbon medium chain triglycerides. PLoS One 2016; 11: e0160159.

104. Uliano A, Stanco M, Lerro M. Perception is not reality: Uncovering the adherence to the Mediterranean diet. J Agric Food Res 2024; 16: 101200.

105. Gouin J-P, Dymarski M. Couples-based health behavior change interventions: A relationship science perspective on the unique opportunities and challenges to improve dyadic health. Compr Psychoneuroendocrinol 2024; 19: 100250.

106. Barnes LL, Dhana K, Liu X, et al. Trial of the MIND diet for prevention of cognitive decline in older persons. N Engl J Med 2023; 389: 602–611.

107. Leta V, Klingelhoefer L, Longardner K, et al. Gastrointestinal barriers to levodopa transport and absorption in Parkinson’s disease. Eur J Neurol 2023; 30: 1465–1480.

108. Staudacher HM, Yao CK, Chey WD, et al. Optimal design of clinical trials of dietary interventions in disorders of gut-brain interaction. Am J Gastroenterol 2022; 117: 973–984.

109. Aarsland D, Batzu L, Halliday GM, et al. Parkinson disease-associated cognitive impairment. Nat Rev Dis Primers 2021; 7: 47.

