## Supplemental File 2 for "A Randomized Safety and Feasibility Crossover Trial of two Mediterranean-Ketogenic Interventions in Individuals with Parkinson’s Disease"

BC Brain Wellness Program

### Modified Mediterranean-Ketogenic Diet

#### What is a Ketogenic (Keto) Diet?

- Ketogenic diets are **high in fat, adequate in protein, and very low in carbohydrates**. Unlike most of the traditional dietary patterns, KD uses fat as the main source of energy instead of carbohydrate. As the utilization of carbohydrates in patients with PD is disturbed, the KD has the potential to restore brain energy deficits.
- On a ketogenic diet, foods high in fat, such as **olive oil, fatty meat, nuts, seeds, and avocado**, are encouraged while sources of carbohydrates, such as grains, legumes, starchy vegetables, and fruits, are restricted or limited.

#### Why a Keto diet?

Ketogenic interventions have been successfully used for decades in epilepsy and more recently in diabetes, obesity, and neurological disorders. Pilot trials of KDs in PD show the possibility of improvement in motor, non-motor, and cognitive symptoms.

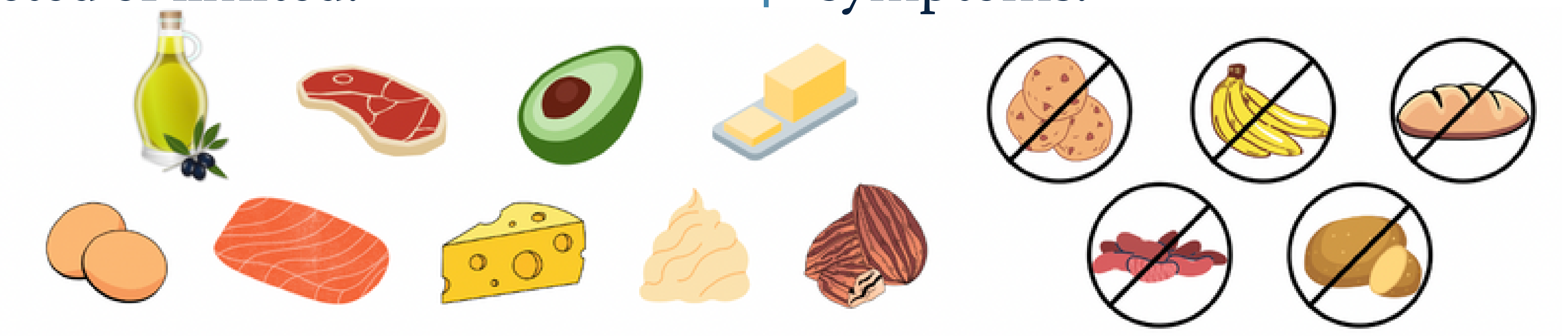

#### What is a Mediterranean (Medi) diet?

The Mediterranean diet is inspired by the dietary pattern of people living in Greece, Spain, and other countries bordering the Mediterranean Sea. The diet features **a high intake of vegetables and fruits, olive oil, nuts, and whole grains**; **a moderate intake of fish and poultry**; and **a low intake of red meat, highly processed food, and sweets.**

#### Why a Medi diet?

Adherence to Mediterranean diets is associated with a reduced risk of developing parkinsonian symptoms and higher age of PD onset.

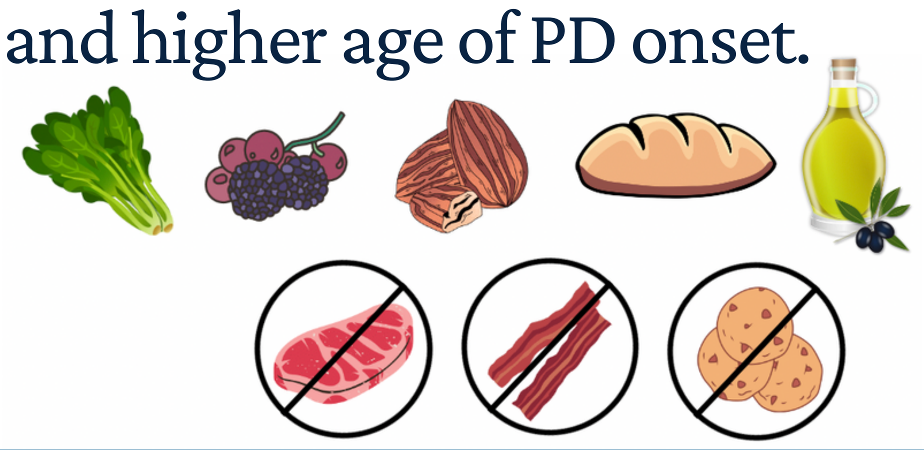

#### What is a Modified Mediterranean-Ketogenic Diet (Medi-KD)?

It is a combination of the Medi diet and the Keto diet. The keto component of the diet will require you to **limit your intake of carbohydrates**, such as grain products and fruits, and **maintain your intake of protein**, such as lean meat and alternatives, while obtaining **most of your energy from fats**. The Mediterranean component of the diet will encourage you to consume more **green leafy vegetables, healthy fats, such as those from nuts and olive oil, and healthy protein sources,** and limit the consumption of processed or fried food, red meat, full-fat dairy, and sweets.

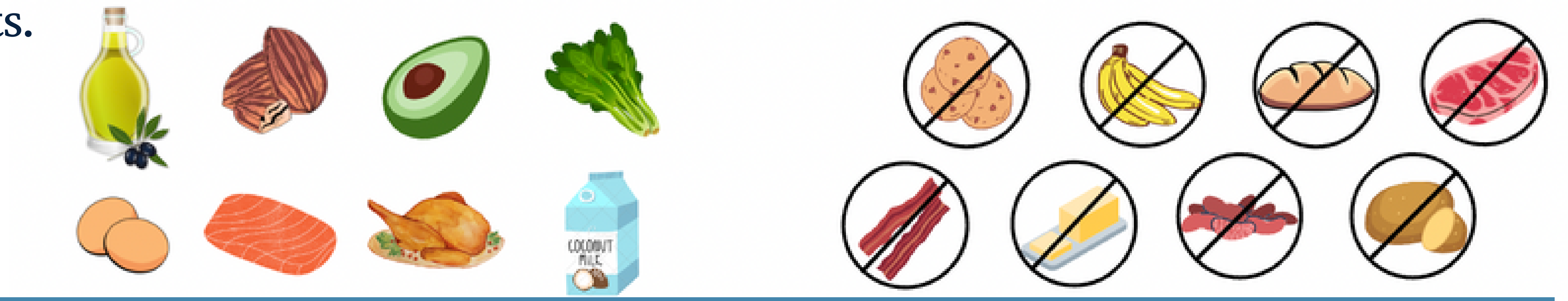

### Modified Mediterranean-Ketogenic Diet

#### Understand Macronutrients

##### 1. Carbohydrates (sugar)

- Carbohydrates, carbs, or sugars, are the primary source of energy in a traditional Western diet. It can be found in a variety of food groups: **grains, fruits, starchy vegetables, legumes, and dairy**. The carbohydrates in these foods are digested and absorbed in our bodies and can quickly turn into fuel for our cells, such as muscle cells and nerve cells.
- Our bodies **preferentially use carbohydrates (glucose) as our energy source**. Therefore, during the Keto diet, when we are training our bodies to use fat as the primary source of energy, we need to restrict the intake of carbohydrates.
- The goal of Medi-KD is to get the allowed carbohydrate intake mostly from non-starchy vegetables.

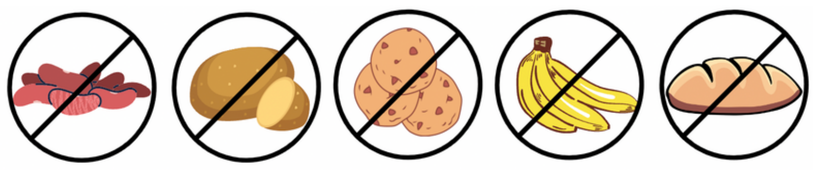

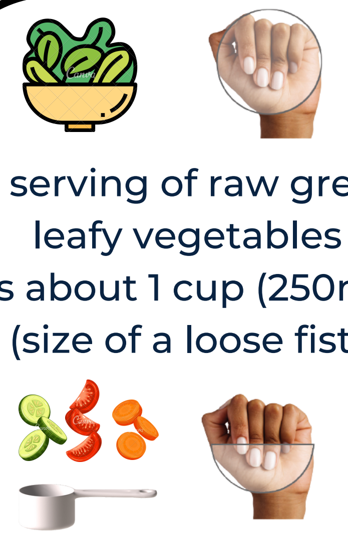

A serving of raw green leafy vegetables is about 1 cup (250ml) (size of a loose fist)

A serving of other vegetables is about 1/2 cup (125ml, size of half a loose fist)

**Non-starchy vegetables**

Eat at least 2 servings of vegetables per day.
One serving is eaten raw or as salad.
Examples: Kale, collards, arugula, spinach, lettuce, peppers, broccoli, celery, green beans, tomatoes, mushroom, zucchini, eggplant

##### 2. Proteins

- Proteins are made up of chemical 'building blocks' called **amino acids.**
- Our bodies use amino acids to **build and repair muscles and bones and to make hormones and enzymes**. They can also be **turned into carbohydrates (glucose)** and used as an energy source. Adequate protein intake is essential for us to stay strong and healthy. However, during the keto diet, we should aim for **a moderate intake of protein**, as excessive protein can be turned into glucose and interrupt our bodies' utilization of fat and the production of ketone bodies (Ketosis).
- Healthy protein products that are encouraged in Mediterranean diets are **fish, poultry, nuts, and seeds.**

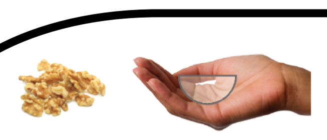

A serving of nuts is 1/4 cup (60ml, a cupped hand)

**Nuts and Seeds**

Eat 3 or more servings of nuts per week
Examples: almonds, hazelnuts, pine nuts, pistachios, walnuts, pecans, cashew

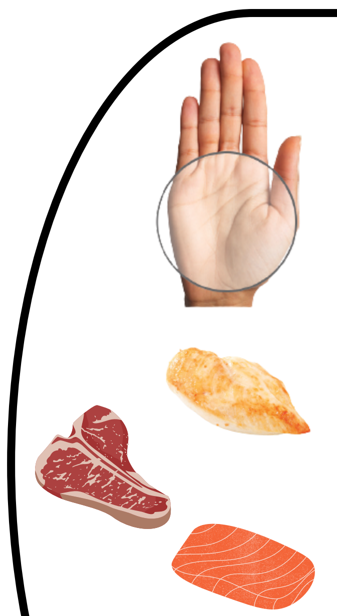

A serving of meat is about 3-5 oz (size of your palm)

**Fish**

Eat 3 or more servings of fish/shellfish per week.
Examples: Salmon, mackerel, herring, rainbow trout, sardines, tuna - skipjack/light

**Poultry**

Preferentially consume poultry than red meat.
Examples: Chicken, turkey, geese, duck, squab

**Red Meats**

Eat less than 1 serving of red or processed meats per day.
Examples: pork, beef, lamb, deli meat (such as roast beef, ham, salami, bologna)

##### 3. Fat (and oil)

- Fats **provide energy** to our bodies, **form the membrane of cells**, and help our bodies **absorb important vitamins**, such as vitamins A, D, E, and K, that are only soluble in fats. Fats also **make foods more flavorful** and **help us feel full**.
- When there is very little intake of carbohydrates and protein, our bodies use fat as the main source of energy. Fats are **broken down into ketone bodies**, which are then taken up by the cells as fuel. For people living with Parkinson's Disease, some of the nerve cells in the brain have trouble utilizing glucose (from carbs) effectively. Therefore, it might be beneficial to switch the energy system to depend on ketone bodies (from fat).
- The Mediterranean diet encourages the intake of **healthy fat sources** that contain mainly **unsaturated fats** that lower your risk of heart disease or are high in **essential fatty acids (omega-3, omega-6)**. Some of the healthy fat sources are nuts, seeds, avocados, fatty fish, vegetable oils, soybeans and soy products, such as tofu.

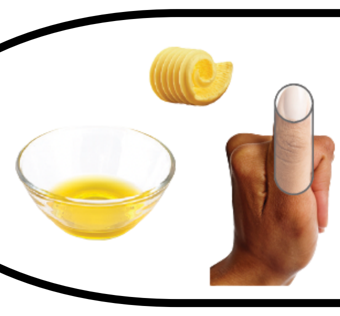

A serving of fat/oil is about 1 tablespoon (15ml, size of a thumb)

**Olive Oil**

Use virgin or extra virgin olive oil as the main source of oil in your diet.
Aim for at least 4 tablespoons per day.

**Butter, Margarine**

Eat less than 1 tablespoon of butter/margarine per day.

### Modified Mediterranean-Ketogenic Diet

#### Possible Side Effects and How to Prevent Them

| | | |
|---|---|---|
| **Dehydration & electrolyte imbalance** | Fatigue, headache, muscle cramping, irritability, difficulty focusing, and heart palpitations, as there is not enough blood volume flowing into your system. This is often referred to as the “keto flu”. | • Try to include **at least 8-12 cups of water per day (2-3L)**<br>• Liberalize salt in your diet (if no hypertension)<br>• Have **1-2 cups of chicken or vegetable broth** or **zero sugar electronic supplement/sports drink per day** for both fluid and electrolyte intake |
| **Constipation** | Changes of bowel habit - difficulty passing the stool, and less frequent bowel movements | • At least **8-12 cups of water per day (2-3L)**<br>• Eat **food high in fibre**: vegetables, nuts, and seeds (especially chia seeds and flaxseeds)<br>• Adequate **physical activities** if possible<br>• Fibre supplement or laxatives (ask dietitian) |
| **Hypotension (low blood pressure)** | Blurred or fading vision, dizziness or lightheadedness, fainting, fatigue, trouble concentrating, nausea | This could be due to **dehydration** as mentioned above. Make sure you have **plenty of oral rehydration solution** (broth, zero-sugar sports drinks)<br>However, **if you are taking medication for blood pressure**, please **report these symptoms to our research coordinator**. A change in the medication dose is needed |
| **Hypoglycemia (low blood sugar)** | Fast heartbeat, shaking, sweating, nervousness or anxiety, irritability or confusion, dizziness, hunger | If you have diabetes, please check your blood sugar and if it's **2.2mmol/L** or less, **have 125mL of orange or apple juice**. Measure blood sugar in 30 minutes after giving the juice. If the blood sugar is still 2.2mmol/L or less give another 30mL of juice and measure the blood sugar in 30 minutes.<br>Please **report these symptoms to our research coordinator.** A change in the medication dose is needed. |

#### What Else Might Impact my Ketone Level?

- **Physical Activities**

When our bodies are in ketosis, they will break down fats and produce ketone bodies for fuel during exercises. Your blood ketone might be elevated if you test it after exercise.
*During the first week of the Keto diet, you might have to limit the amount of physical activity as your body is transitioning fuels and might not be ready to provide enough for strenuous activity. You may need to slowly build your exercise tolerance while your body adapts to using ketones for fuel. Once it is adapted, you should be able to exercise for even longer periods than when you depended on glucose for fuel!

- **Sickness**

Before and during sick days, the hormones in your body will fluctuate and might cause your ketone level to drop. It is important to maintain your energy and fluid intake during sick times. You may choose more blenderized meals or stew to replenish energy and fluid at the same time. Taking oral rehydration solutions, such as Powerade Zero, diet ginger ale, or chicken/vegetable broth is a good way to ensure enough fluid and electrolyte intake.
*Make sure you document the days that you get sick and report to our dietitian at your weekly check-in.

### Modified Mediterranean-Ketogenic Diet

#### Medi-KD Timeline

| | |
|---|---|
| **Pre-intervention visit (today)** | Learn to use Keto Diet Calculator and make your own menu;<br>Start Keto Ratio 1:1 (1 part fat to 1 part the sum of protein and carb) |
| **Week 1 Follow Up (3-4 days from now)** | **The day before follow-up:**<br>☐ 1-day Food Diary<br>☐ Test blood ketone before bed<br>**After Follow-Up:**<br>Increase Keto Ratio to 1.5:1 (1.5 part fat to 1 part sum of protein and carb) |
| **Week 2 Follow Up** | **The day before follow-up:**<br>☐ 1-day Food Diary<br>☐ Test blood ketone before bed<br>**After Follow-Up:**<br>Increase Keto Ratio to 2:1 (2 part fat to 1 part sum of protein and carb) |
| **6 More Weekly Follow-Ups** | **Choose 1 day of the week before the follow-up:**<br>☐ 1-day Food Diary<br>☐ Test blood ketone before bed<br>**During the follow-up:**<br>Ask questions and troubleshoot |
| **Post-intervention visit** | Bring back the ketone test kit and study journal;<br>Complete questionnaires and clinic measurements |

#### Who to Contact

You can first record any questions in the study journal and discuss them with our dietitian at the weekly check-in. Our dietitian will direct you to a clinician if the question stays unresolved.

If your questions are urgent, please contact our research coordinator at 604-827-1950.

If it is an emergency, please call 604-822-7121 UBC Hospital Switchboard and ask for Dr. Silke Appel-Cresswell.

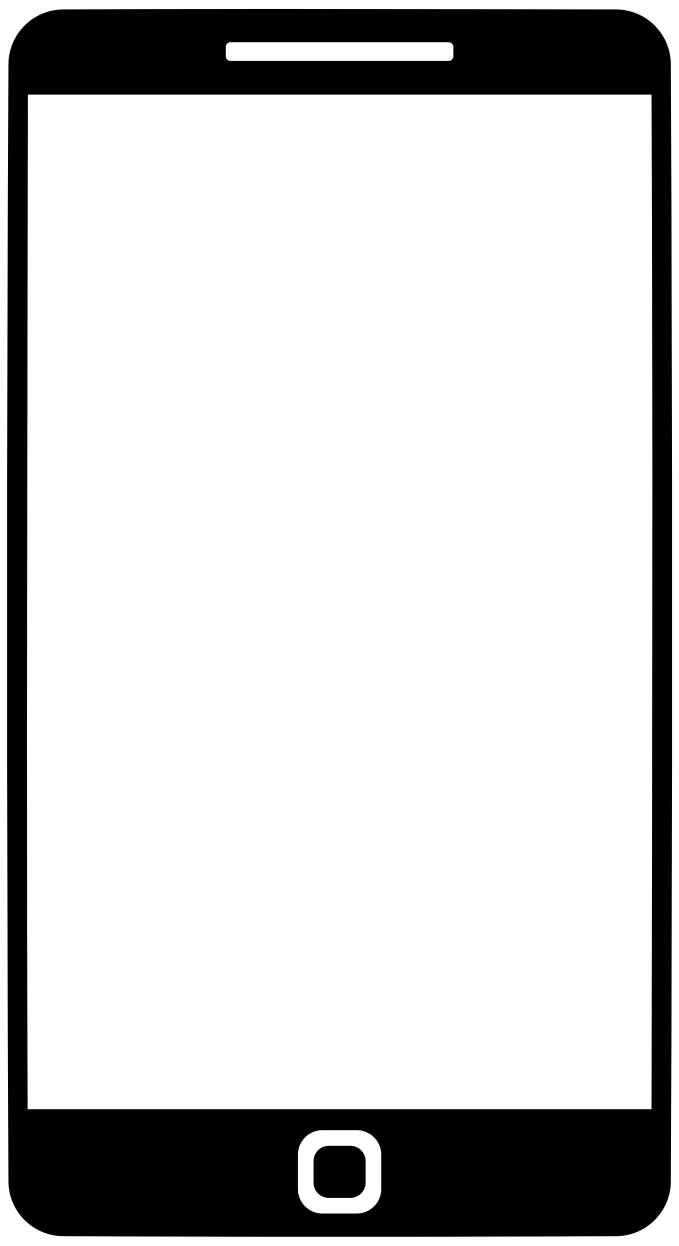

**BC Brain Wellness Program**
***Brain Wellness, Beyond All Boundaries.***

### Mediterranean Diet + MCT Oil

#### What is a Mediterranean (Medi) diet?

The Mediterranean diet is inspired by the dietary pattern of people living in Greece, Spain, and other countries bordering the Mediterranean Sea. The diet features **a high intake of vegetables and fruits, olive oil, nuts, and whole grains**; **a moderate intake of fish and poultry**; and **a low intake of red meat, highly processed food, and sweets.**

#### Why a Medi diet?

Adherence to Mediterranean diets is associated with a reduced risk of developing parkinsonian symptoms and higher age of PD onset.

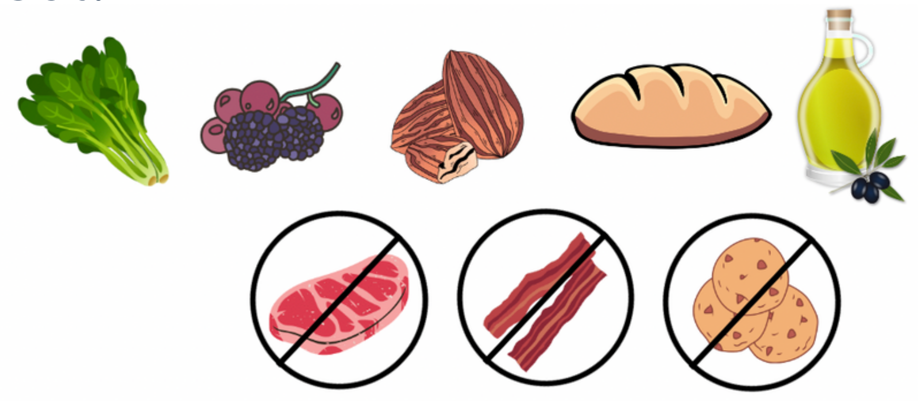

#### What is MCT Oil?

- MCT oil is short for **Medium Chain Triglycerides**.
- It is **a type of fat** that our body can easily absorb.
- Once absorbed, the medium-chain fatty acids are directly metabolized by our liver, where they will be broken down into **ketone bodies** and released into our bloodstream.

#### Why MCT Oil?

- When we are consuming a typical Western diet, carbohydrates (sugars, complex or simple) make up the majority of our energy intake. All other types of carbohydrates will be transformed into glucose before they are directly used by all cells in our bodies.
- For people living with Parkinson's disease, some brain cells cannot effectively use glucose for energy. The **ketone bodies**, metabolized from MCT oil, can **serve as an alternative energy source** and are theorized to sustain the functions of those brain cells.

#### What does Mediterranean-MCT diet look like?

- We provide __ Mediterranean recipes that you can choose from. The following pages will introduce you to the main components of the Mediterranean diet. You can also work with our dietitian to create your own recipes once you have a better idea of what the diet is like.
- The MCT oil will contribute to part of your calorie intake, so the total amount of food you eat might be slightly less than your regular intake. However, for people who would benefit from weight gain, the amount of food does not need to be changed or might need to be increased. Your dietitian will discuss this further with you.

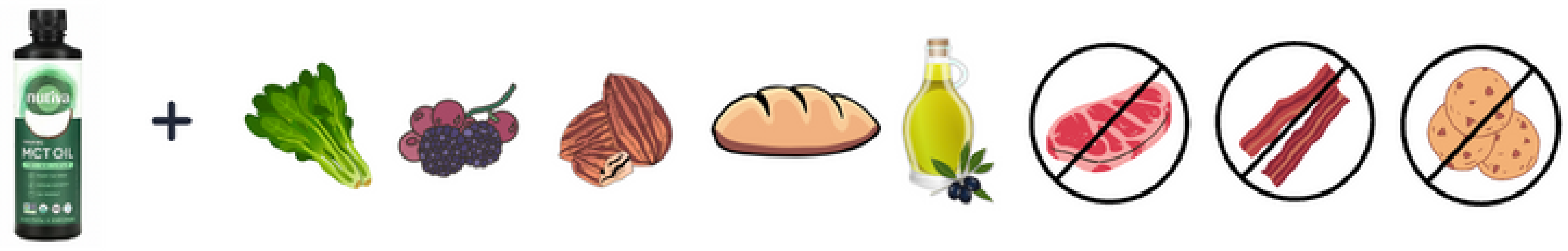

BC
Brain
Wellness
Program

### Mediterranean Diet + MCT Oil

#### 1. Fruits and Vegetables

##### Vegetables

Eat at least 2 servings of vegetables per day.
At least one of the servings is eaten as salad or raw.

Examples of green leafy vegetables: Kale, collards, arugula, salad greens, spinach, lettuce, watercress

Examples of other vegetables: peppers, squash, carrots, broccoli, celery, green beans, potatoes, tomatoes, beets, zucchini, eggplant

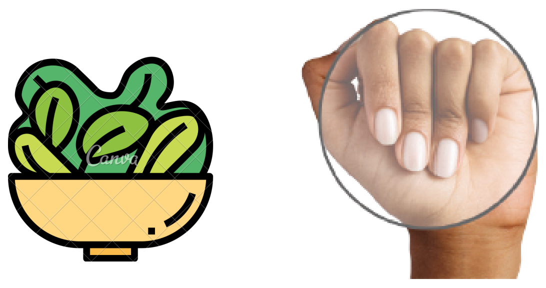

A serving of raw green leafy vegetables is about 1 cup (250ml) (size of a loose fist)

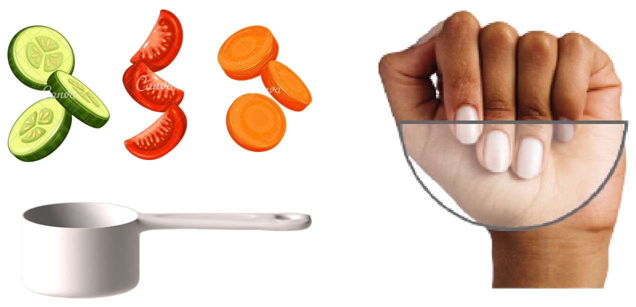

A serving of cooked green leafy vegetables or other vegetables is about 1/2 cup (125ml) (size of half a loose fist)

##### Fruits

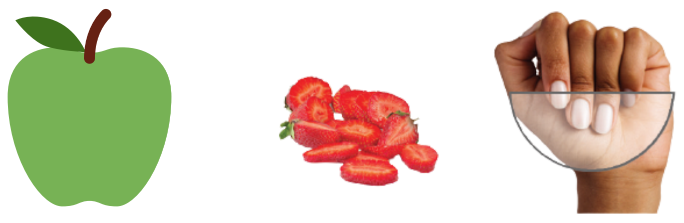

A serving of fruit is about 1/2 cup (125ml, the size of half a loose fist), or one whole fruit.

Eat at least 3 servings of fruits per day.

Examples: Strawberries, blueberries, raspberries, blackberries, apples, apricots, pears

#### 2. Sauce

##### Sofrito

Consume 2 or more times of dishes seasoned with Sofrito per week.

Sofrito: sauce made with tomato and onion, leek, or garlic, simmered with olive oil

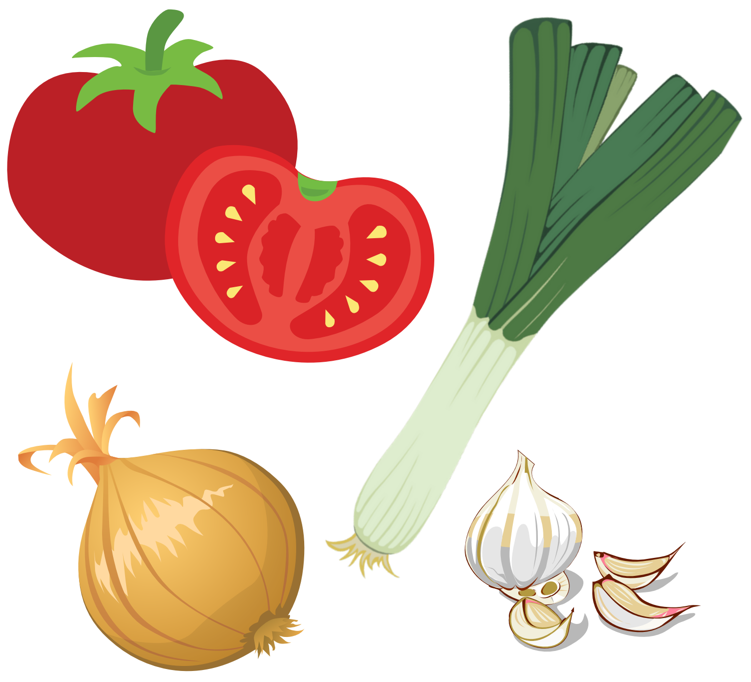

#### 3. Protein - Plant

##### Beans*

Eat at least 3 servings of beans and legumes per week.

Examples: kidney beans, red and green lentils, split peas, chickpeas, black beans, navy beans, and black-eye peas.

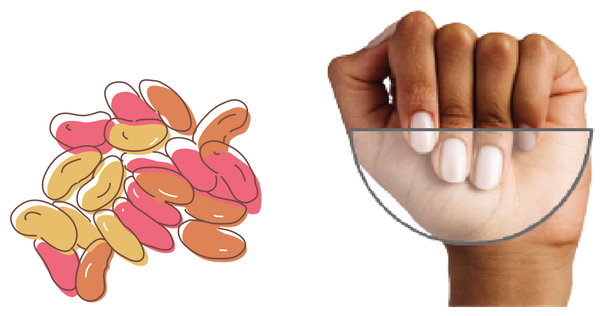

A serving of beans/legumes is about 150g (size of half a loose fist)

##### Nuts*

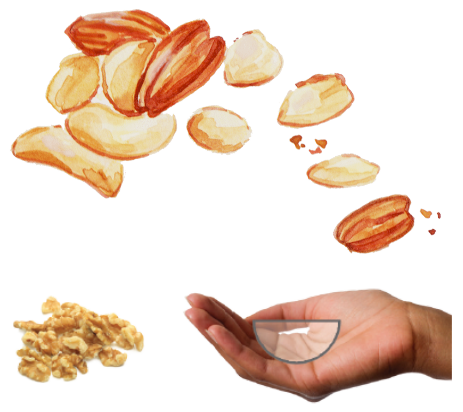

A serving of nuts is 1/4 cup (30g, a cupped hand)

Eat 3 or more servings of nuts per week
Examples: almonds, hazelnuts, pine nuts, pistachios, and walnuts, pecans, cashew

*Protein from Plant

Natural plant-based proteins tend to have a good amount of dietary fiber, vitamins, minerals, and phytochemicals.

Choosing more plant-based proteins is related to decreased risks of

- Heart disease
- High blood pressure (hypertension)
- High cholesterol
- Many cancers
- Obesity
- Stroke
- Type 2 diabetes

BC Brain Wellness Program

### Mediterranean Diet + MCT Oil

#### 4. Protein - Animal

##### Fish/Shellfish

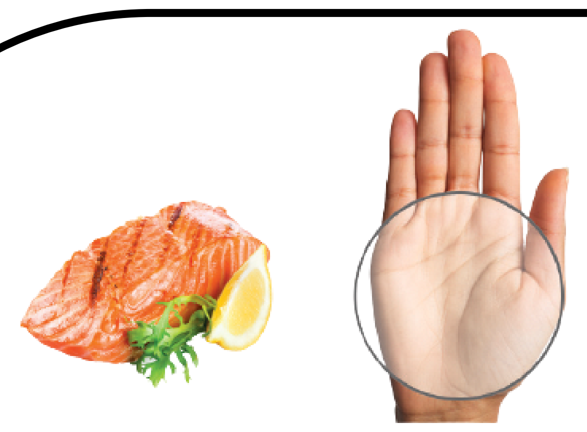

A serving of fish is about 100-150g (the size of your palm)

Eat 3 or more servings of fish or shellfish per week.

Examples: Salmon, mackerel, herring, rainbow trout, sardine, tuna skipjack/light, mussel, oyster, scallop

##### Poultry

Choose poultry over red meat.

Examples:
Chicken, turkey, geese, duck, squab

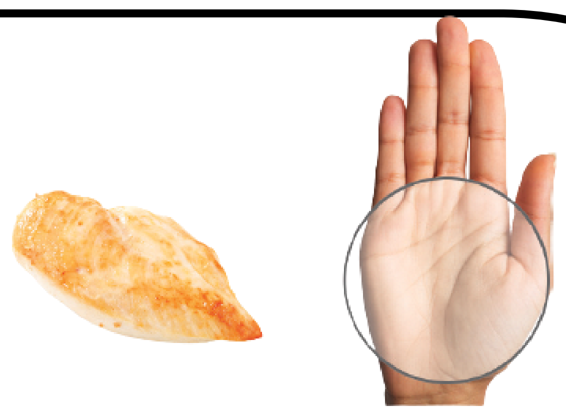

A serving of poultry is about 100-150g (size of your palm)

##### Red Meat

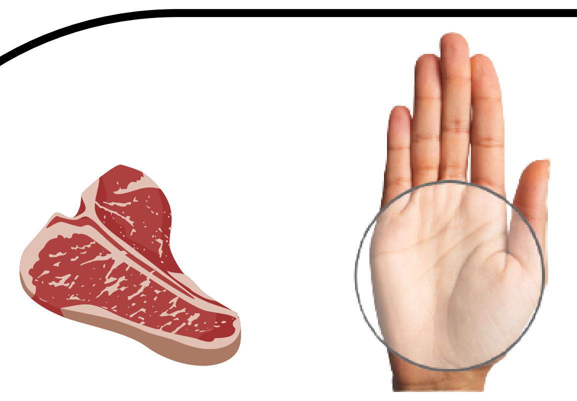

A serving of red meat is about 100-150g (size of your palm)

Eat less than 1 serving of red or processed meats per day.

Examples: pork, beef, lamb, deli meat (such as roast beef, ham, salami, bologna)

#### 5. Oil

##### Olive Oil

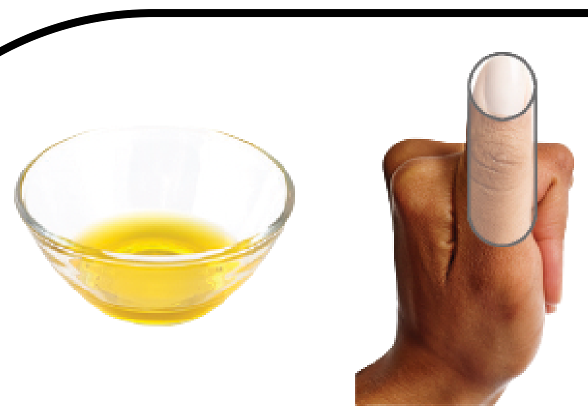

A serving of olive oil is about 1 tablespoon (15ml) (size of a thumb)

Use virgin or extra virgin olive oil as the main culinary fat.

Aim for at least 4 tablespoons per day.

##### Butter, Margarine

Eat less than 1 tablespoon of butter/margarine/cream per day.

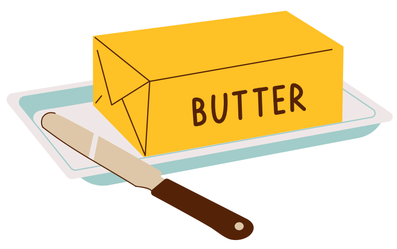


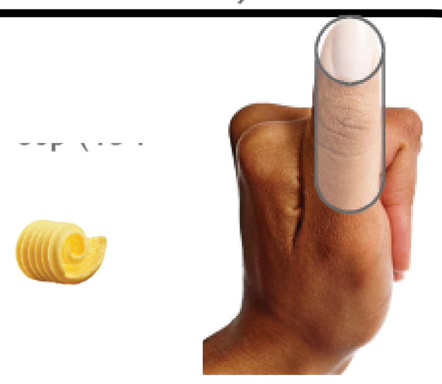

A serving of butter/margarine/cream is about 1 tablespoon (15ml) (size of a thumb)

#### 6. Others

##### Alcohol

Having 1 servings of wine per day if desired.

A serving of wine is 150 ml for men, 100 ml for women

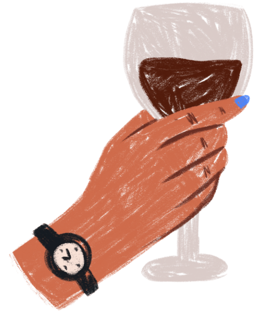

##### Sweet/ carbonated Drink

Have less than 1 serving of sweet or carbonated beverage per day.

Examples: Coke, Sprite, Fanta, orange juice

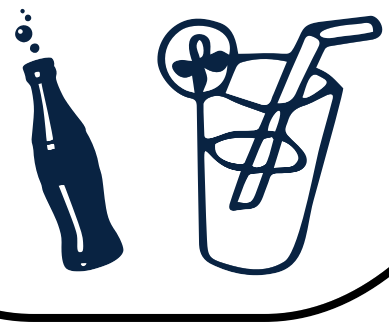

##### Pastries, Sweets

Eat commercial pastries and sweets less than 3 times per week.

Examples: Biscuits/rolls, cake, Danish, sweet rolls, donuts, cookies, brownies, pie, candy bars, ice cream, pudding, milkshakes/frappes

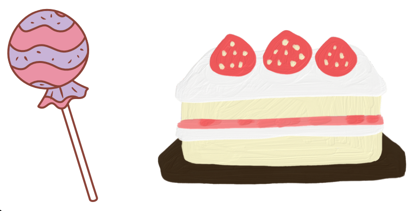

BC Brain Wellness Program

### Mediterranean Diet + MCT Oil

#### Possible Side Effects and How to Prevent Them

All drugs may cause side effects. However, many people have no side effects or only minor side effects. Some possible side effects of MCT oil include:

- Belly pain
- Nausea
- Headache
- Diarrhea
- Upset stomach or throwing up

Our study design tries to reduce the risk of side effects by slowly increasing the MCT oil dose, so your body will have plenty of time to adapt to it. Taking the MCT oil with food might also alleviate the effect.

Please **call your research coordinator at 604-827-4230 or get medical help** if any of the side effects bother you or do not go away, or **if you have any of the symptoms listed below**:

- Signs of an allergic reaction, like rash; hives; itching; red, swollen, blistered, or peeling skin with or without fever; wheezing; tightness in the chest or throat; trouble breathing, swallowing, or talking; unusual hoarseness; or swelling of the mouth, face, lips, tongue, or throat
- Very upset stomach or throwing up
- Very bad belly pain
- Very bad skin irritation

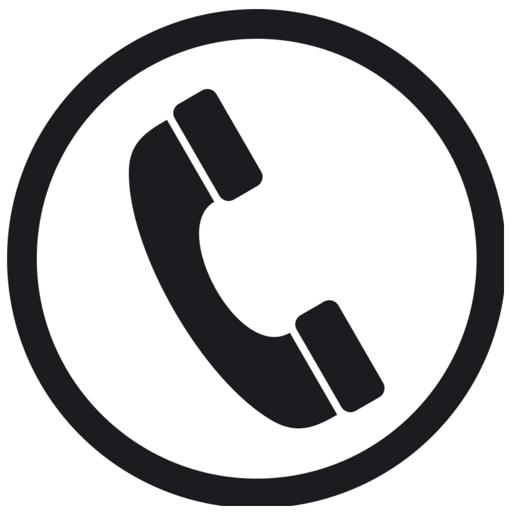

#### What to do if I miss a dose?

- Take a missed dose as soon as you think about it.
- If it is close to the time for your next dose (less than 3 hours), skip the missed dose and go back to your normal time.
- Do not take 2 doses at the same time or extra doses.

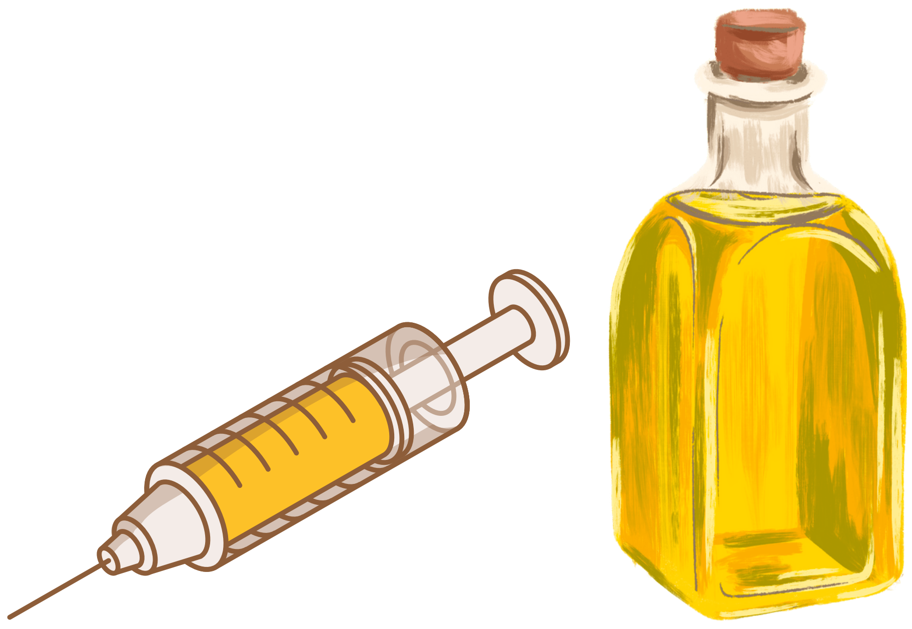

BC Brain Wellness Program

### Mediterranean Diet + MCT Oil

#### Medi-MCT Timeline

**Pre-intervention visit (today)**

Learn to use the Cronometer app to look up the recipes and make your own menu;
Start MCT oil 5ml per dose, 2 doses per day

**Week 1 Follow Up (3-4 days from now)**

**The day before follow-up:**

- ☐ 1-day Food Diary
- ☐ Test blood ketones before bed

**After Follow-Up:**

Increase MCT oil to 10ml per dose, 2 doses per day

**Week 2 Follow Up**

**The day before follow-up:**

- ☐ 1-day Food Diary
- ☐ Test blood ketones before bed

**After Follow-Up:**

Increase MCT oil to 15ml per dose, 2 doses per day

**6 More Weekly Follow-Ups**

**Choose 1 day of the week before the follow-up:**

- ☐ 1-day Food Diary
- ☐ Test blood ketones before bed

**During the follow-up:**

Ask questions and troubleshoot

**Post-intervention visit**

Bring back the ketone test kit, MCT oil, and study journal;
Complete questionnaires and clinic measurements

#### Who to Contact

You can first record any questions in the study journal and discuss them with our dietitian at the weekly check-in. Our dietitian will direct you to a clinician if the question stays unresolved.
If your questions are urgent, please contact our research coordinator at 604-827-1905.
If it is an emergency, please call 604-822-7121 UBC Hospital Switchboard and ask for Dr. Silke Appel-Cresswell.

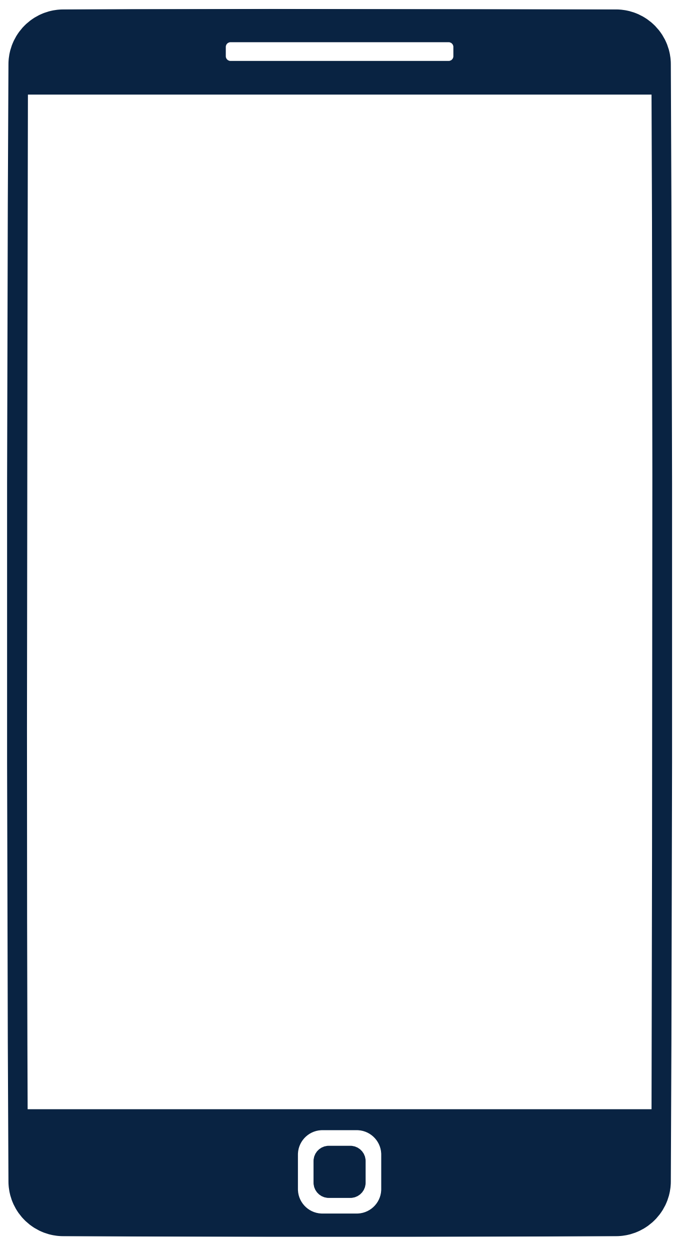

**BC Brain Wellness Program**
***Brain Wellness, Beyond All Boundaries.***
