## Supplemental File 1 for "A Randomized Safety and Feasibility Crossover Trial of two Mediterranean-Ketogenic Interventions in Individuals with Parkinson’s Disease"

**Supplemental Table 1. Eligibility Criteria**

| **Inclusion Criteria** | **Exclusion Criteria** |
| --- | --- |
| Age 40-85 years | Atypical parkinsonism |
| PD diagnosis based on Movement Disorder Society (MDS) criteria.^62^ | Medical or psychiatric condition that would prevent full study participation |
| Hoehn & Yahr stage 1-3 | Pregnancy |
| On stable dose of dopaminergic medication for at least one month (including no medication where applicable) | Significant dysphagia |
|  | Diabetes on insulin |
|  | Warfarin use |
|  | Inflammatory bowel disease |
|  | Dementia defined by a MoCA score <21 |
|  | Inability to fill in electronic questionnaires or understand study instructions |
|  | Use of immunomodulatory agents |
|  | Probiotic use in the previous 4 weeks (except for dietary sources such as yoghurt, kefir etc.), or antibiotic use in the 3 months prior to starting the trial |
|  | Use of MCT oil or adherence to a ketogenic diet in the 8 weeks prior to the trial |
|  | Allergy to MCT oil, coconut oil, or coconut |

**Supplemental Table 2. Pre- and Post-Intervention MEDAS Adherence Scores.**

|  | **First baseline  N = 52^1^** | **Second baseline  N = 40^1^** | **MeDi-KD  N = 39^1^** | **MeDi-MCT  N = 41^1^** |
| --- | --- | --- | --- | --- |
| **MEDAS Category** |  |  |  |  |
| **Good to very good adherence** | 0 (0%) | 1 (2.8%) | 1 (3.0%) | 8 (23%) |
| **Moderate to fair adherence** | 22 (48%) | 21 (58%) | 25 (76%) | 19 (54%) |
| **Weak adherence** | 24 (52%) | 14 (39%) | 7 (21%) | 8 (23%) |
| **^1^n (%)** | | | | |
